# Recalibrating Mendelian randomization under winner’s curse, sample structure and polygenicity

**DOI:** 10.64898/2026.06.25.26356593

**Authors:** Yihe Yang, Zhaotong Lin, Haoran Xue, Xiaofeng Zhu

## Abstract

Recently, Hu et al. (2024) conducted a benchmarking study showing that most existing Mendelian randomization (MR) methods exhibit substantial bias and inflated type-I error rates in real data. They attributed these failures to two largely neglected sources of bias: winner’s curse and polygenicity-induced bias. Although a few methods have been developed to address one or both of these issues, existing approaches either do not fully account for both biases or are restricted to the univariable setting. In this paper, we propose a multivariable Rao-Blackwellization that corrects winner’s curse while accounting for polygenicity and sample structure in a unified framework. Unlike univariable Rao-Blackwellization, where instrument selection yields a truncated normal statistic amenable to a Mills-ratio correction, multivariable Rao-Blackwellization conditions on a noncentral *χ*^2^ statistic, for which no analogous correction is available. We derive closed-form conditional moments under this instrument selection model and use them to construct bias-corrected summary statistics that can be integrated into a wide range of existing MR methods. Simulations and real data analyses show that, when combined with methods such as MR-cML and MR-BEE, the proposed correction substantially improves type-I error control and yields more robust inference.

## 1 Introduction

Mendelian randomization (MR) is a powerful tool that uses genetic variants as instrumental variables (IVs) to infer the causal effects of exposures on outcomes. Because summary statistics of large-scale genome-wide association studies (GWAS) on various complex traits and diseases have become increasingly available, MR has been widely applied in the past decade. To ensure unbiased inference of the causal effect, MR relies on three core assumptions, known as the valid IV assumptions: the genetic variants must (i) be associated with the exposure, (ii) contribute to the outcome only through the exposure, and (iii) be independent of all confounders^[1]^. In practice, these assumptions are often violated and are difficult to verify. Horizontal pleiotropy is one major challenge, where genetic variants may affect the outcome through pathways other than the target exposures. Weak genetic associations can induce weak IV bias, which can be further amplified by overlap between the GWAS samples for the exposure and the outcome^[2]^. Many MR methods have been developed to address these challenges.

Despite the proliferation of MR methods, a recent benchmarking study^[3]^ raised serious concerns about their reliability in inferring and estimating causal effects in real-world settings. In their positive control analysis, the causal effect of a trait on itself should be equal to one. However, most of the existing methods produced attenuated estimates. This attenuation is primarily driven by winner’s curse, where selecting variants based on statistical significance leads to overestimation of genetic variant effects on the exposure, thereby biasing the causal estimate toward the null. In their negative control analyses using trait pairs with no plausible causal relationship, most of the existing methods failed to control type-I error rates under real-data confounding scenarios involving population stratification (manifesting as inflation of LDSC intercepts), polygenicity (manifesting as non-zero genetic correlation between trait pairs), and family-level confounders. Among the benchmarked methods, MR-APSS is the only one that maintained robust statistical inference in both positive and negative control analyses by explicitly modeling winner’s curse, the polygenic background, and sample structure, within a likelihood framework^[4]^. However, MR-APSS is a univariable MR method, and its framework cannot be directly incorporated into other existing popular MR methods. A general correction framework that extends to MVMR and integrates with existing MR methods is still lacking.

Randomized IV selection provides a promising route toward such a framework. Ma et al.^[5]^ showed that, by introducing auxiliary noise at the IV selection stage and then applying Rao–Blackwellization, one can construct adjusted estimates of genetic effects on exposure that are independent of the selection event, thereby effectively removing the winner’s curse bias. Xie et al.^[6]^ later built on this idea in MR-CARE to improve robustness to horizontal pleiotropy. However, both Ma et al.^[5]^ and Xie et al.^[6]^ focused on UVMR and addressed only the winner’s curse, leaving bias from polygenicity and sample structure uncorrected. In particular, extending this framework to MVMR is also nontrivial, because joint IV selection across multiple exposures induces a multivariate truncation region that cannot be addressed by the Mills-ratio correction used in the univariate case.

To address these gaps, we develop a multivariable Rao–Blackwellization that simultaneously corrects winner’s curse and bias arising from polygenicity and sample structure. The proposed framework extends randomized-selection correction from UVMR to MVMR and jointly models the exposure and outcome summary statistics, thereby accommodating sample overlap. Unlike UVMR, where IV selection induces a truncated-normal conditional distribution, joint IV selection in MVMR leads to a noncentral *χ*^2^ conditional distribution and therefore requires new conditional-moment calculations. We derive closed-form conditional moments for this setting using the Poisson mixture representation of the noncentral *χ*^2^ distribution and score functions based on derivatives of its log-survival function, while accounting for polygenicity and sample structure. This correction yields adjusted summary statistics and covariance estimates, which can be used as a plug-in correction layer for existing MR procedures. We illustrate this by pairing the correction with MR-cML^[7,8]^ and MR-BEE^[9]^, which we name MR-cML-RB and MR-BEE-RB, respectively.

We show in the simulations, positive control benchmark, and negative control benchmark analyses that MR-cML-RB and MR-BEE-RB substantially improve calibration and robustness over MR-APSS, MR-CARE, and other UVMR methods. We further apply MR-cML-RB and MR-BEE-RB to an MVMR analysis investigating the causal contributions of multiple common risk factors to coronary artery disease (CAD), including low-density lipoprotein (LDL) cholesterol, high-density lipoprotein (HDL) cholesterol, and type 2 diabetes (T2D).

After accounting for polygenicity, sample structure and winner’s curse, the previously reported protective effect of HDL cholesterol on CAD is no longer significant. This result may explain a long-standing inconsistency in the MR literature: HDL cholesterol has frequently been reported to have a significant causal effect on CAD, although this association has been considered biologically questionable and is not supported by therapeutic evidence^[9,10,11]^.

## 2 Method

### 2.1 Mendelian randomization model under polygenicity

Our method takes GWAS summary statistics as input and explicitly decomposes them into a foreground signal and background noise that includes polygenicity, unlike classical MR methods that typically treat the background as GWAS estimation error only. Suppose we have *m* independent genetic variants and *p* exposures. For the *j*th genetic variant, let 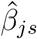 and se 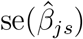denote the GWAS effect estimate and its standard error for the *s*th exposure, and let 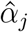 and se 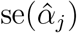 denote the corresponding quantities for the outcome. We write 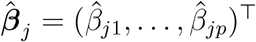 and 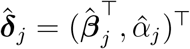, and assume that the GWAS summary statistics are computed on standardized phenotypes and genotypes. Let ***θ*** = (*θ*_1_, …, *θ*_*p*_)^⊤^ denote the causal effects of the exposures on the outcome, ***β***_*j*_ = (*β*_*j*1_, …, *β*_*jp*_)^⊤^ denote the unknown fixed effects of variant *j* on the exposures, *γ*_*j*_ denote its pleiotropic effect on the outcome not mediated by the exposures, and 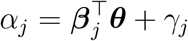 denote the total genetic effect on the outcome. We write 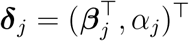 for the vector of true genetic effects. The observed summary statistics for each variant *j* are decomposed as:

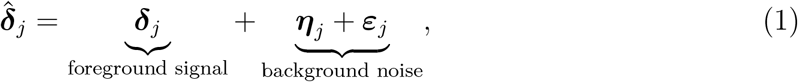

which, unlike the traditional model^[8,9]^, explicitly incorporates polygenicity through the term ***η***_*j*_ in the background noise^[4]^. Specifically, ***η***_*j*_ = ∑_*ℓ*_ *r*_*jℓ*_***φ***_*ℓ*_ is the LD-tagged polygenic component, with *r*_*jℓ*_ being the LD correlation between variants *j* and *ℓ* and ***φ***_*ℓ*_ being the random polygenic effect vector of variant *ℓ* on the exposures and the outcome, and ***ε***_*j*_ is the GWAS estimation error, encompassing finite-sample variability and error covariance induced by sample overlap, population stratification, and cryptic relatedness. Moreover, ***η***_*j*_ and ***ε***_*j*_ are assumed to be independent of each other^[4]^.

For the polygenic component, we assume the polygenic effect ***φ***_*ℓ*_ follows a normal distribution with E(***φ***_*ℓ*_) = **0** and cov(***φ***_*ℓ*_) = **Ω** independently across variants, where

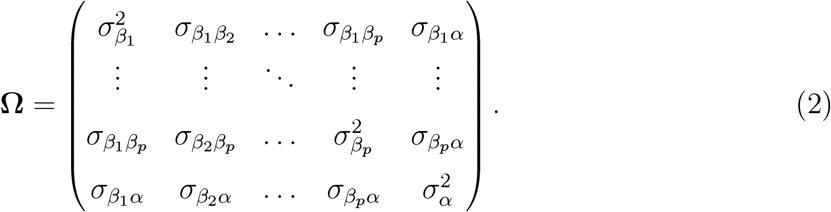

In this covariance matrix, 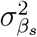 and 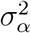 denote the per-SNP variance of polygenic effects on the *s*th exposure and outcome, respectively, and 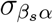 and 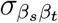 denote the corresponding per-SNP covariances, for 1 ≤ *s < t* ≤ *p*. As a result, the covariance matrix of the LD-tagged polygenic component ***η***_*j*_ is

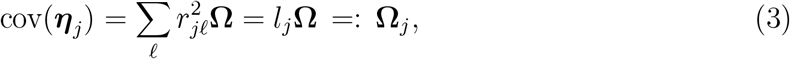

where 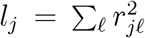 is the LD score of variant *j*. In practice, we estimate **Ω** using the procedure of Hu et al.^[4]^: the diagonal entries 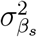 and 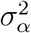 are approximated by the per-SNP heritabilities estimated by univariate LD score regression (LDSC) slope parameters^[12]^, and the off-diagonal entries 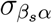 and 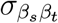 by the per-SNP genetic covariances estimated by bivariate LDSC slope parameters^[13]^.

For the estimation error ***ε***_*j*_ in model (1), we parameterize its covariance by cov(***ε***_*j*_) = **D**_*j*_**R**_***εε***_**D**_*j*_, where 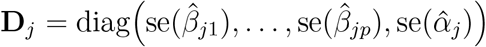 captures the SNP-specific standard errors of the exposure and outcome GWAS estimates, and **R**_***εε***_ captures the sample structure in GWAS data including population stratification, cryptic relatedness, and sample overlap^[4]^. In practice, the diagonal elements of **R**_***εε***_ are estimated by univariate LDSC intercept parameters to correct for population stratification and cryptic relatedness in the GWAS^[12]^. The off-diagonal elements are estimated by bivariate LDSC intercept parameters to capture the covariance among GWAS estimation errors due to sample overlap or population stratification^[13]^.

Under the large-sample theory, the summary statistics are asymptotically normal^[14]^: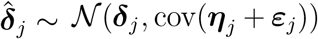. Combining the LD-tagged polygenic component ***η***_*j*_ and the estimation error component ***ε***_*j*_ derived above, their covariances add to give 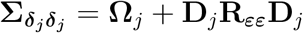, which leads to the working model for the GWAS summary statistics,

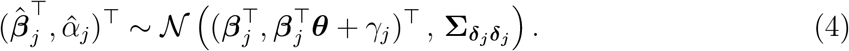

Throughout this work, we treat the two covariance components obtained from LDSC, and thus 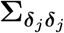, as fixed plug-in quantities.

It should be pointed out that existing MR methods differ in how they specify the GWAS error covariance **R**_***εε***_ and whether they model the polygenic background component **Ω**_*j*_ as summarized in Table 1. The most restrictive methods set **R**_***εε***_ = **I**, assuming no correlation among the exposure and outcome GWAS estimation errors, as would be expected under no sample overlap and no additional GWAS inflation. More recent methods, like Lin et al.^[15]^, Lorincz-Comi et al.^[9]^, accommodate sample overlap by modeling the off-diagonal correlations, but still strictly fix the diagonal entries at one, thereby ignoring residual GWAS inflation from uncontrolled population stratification, cryptic relatedness, and other confounders. Lastly, Hu et al.^[4]^ is the only method that explicitly allows the diagonal entries to exceed one and also models the polygenic background component.

**Table 1:** Summary of selected MR methods and the biases they explicitly address.

| Method | Setting | Weak-IV bias | Horizontal pleiotropy | Winner's curse | Sample overlap | Residual GWAS inflation* | Polygenic background | Note |
| --- | --- | --- | --- | --- | --- | --- | --- | --- |
| IVW <sup>[16]</sup> | UV+MV | – | – | – | – | – | – | Baseline requiring strong, valid IVs. |
| Weighted mode <sup>[17]</sup> | UV | – | ✓ | – | – | – | – | Relies on the zero-modal pleiotropy assumption; typically less precise. |
| MR-APSS <sup>[4]</sup> | UV | ✓ | ✓ | ✓ | ✓ | ✓ | ✓ | Foreground–background likelihood with a foreground InSIDE assumption. |
| RIVW <sup>[5]</sup> | UV | ✓ | – | ✓ | – | – | – | Rerandomized IV selection developed for independent GWASs. |
| MR-cML <sup>[8]</sup> | UV+MV | ✓ | ✓ | – | ✓ | – | – | Pleiotropy robustness under a plurality-validity condition. |
| MR-BEE <sup>[9]</sup> | UV+MV | ✓ | ✓ | – | ✓ | – | – | Bias-corrected estimating equations with iterative pleiotropy handling. |
| MR-CARE <sup>[6]</sup> | UV | ✓ | ✓ | ✓ | – | – | – | Rerandomized selection with robust invalid-IV screening. |
| MR-cML-RB (this paper) | UV+MV | ✓ | ✓ | ✓ | ✓ | ✓ | ✓ | Weak-IV and pleiotropy properties inherited from MR-cML. |
| MR-BEE-RB (this paper) | UV+MV | ✓ | ✓ | ✓ | ✓ | ✓ | ✓ | Weak-IV and pleiotropy properties inherited from MR-BEE. |

\* Residual GWAS inflation refers to diagonal error inflation from sources such as population stratification and cryptic relatedness.

### 2.2 Winner’s curse

In practice, clumping and thresholding (C+T) is typically used to select independent IVs for MR. For UVMR, a variant is selected if its marginal association with the exposure exceeds a significance threshold:

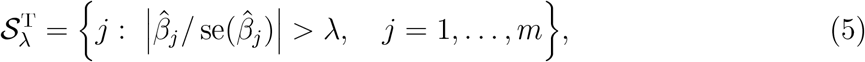

where, for example, *λ* = 5.45 corresponds to the usual GWAS significance level 5 × 10^−8^. For MVMR, the usual IV selection principle is to select variants associated with at least one exposure. Two screening strategies are commonly used in practice. One is a joint test across the *p* exposures, selecting variants with 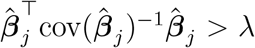, where *λ* is chosen so that 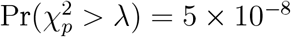^[9]^. The other is to screen each exposure separately using the univariable rule and then take the union of the selected variants across exposures ^[18]^. The thresholding step is then followed by LD clumping, which sorts variants by their association strength and removes all nearby variants in LD with each index variant (e.g., *r*^2^ *>* 0.001), retaining approximately independent IVs.

However, this selection strategy introduces the winner’s curse when the same GWASs are used both to select instruments and to estimate their SNP-exposure effects^[5]^. Among the selected variants, the observed exposure estimates 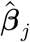 tend to have overestimated exposure associations, especially for those near the significance threshold. In a UVMR setting, the inflated SNP-exposure estimates can attenuate causal effect estimates toward the null. On the other hand, when the exposure and outcome GWASs are correlated, for example, due to sample overlap, the selection will also introduce bias in outcome associations. The bias in the outcome associations may offset the usual attenuation and even produce upward bias^[19,20]^. Although this issue is often overlooked due to the large and continually increasing sample sizes of GWASs, Hu et al.^[3]^ demonstrates that the winner’s curse can still induce substantial bias in current applications.

### 2.3 Multivariable Rao-Blackwellization

We propose a multivariable Rao-Blackwellization to correct winner’s curse while simultaneously accounting for the polygenic background and sample structure in the GWAS summary statistics. Similar to the original Rao-Blackwellization^[5]^, the proposed extension consists of two key steps: re-randomized IV selection and winner’s curse removal via Rao-Blackwellization.

In the first step, we generate an auxiliary random vector

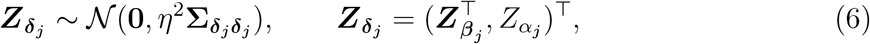

independently of the observed GWAS estimates 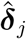, for *j* = 1,…,*m*, where *η* is a tuning parameter. To apply Rao-Blackwellization, an IV will be selected jointly across all exposures via the following *χ*^2^ statistic. Specifically, the *j*th variant is selected as an IV if

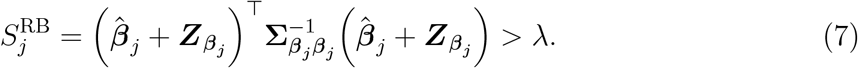

The selected IV set is then defined as 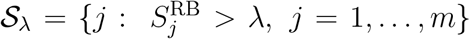. Here 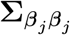 denotes the exposure block of the full covariance 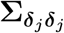 in (4), including both the sampling error and polygenic background. In practice, the threshold *λ* is commonly set as the (1 − 5 × 10^−8^) quantile of a 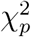 distribution, that is, 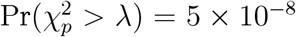. The tuning parameter *η* is typically fixed at *η* = 0.5 or a similar value^[5]^. A first key distinction from Ma et al.^[5]^ is the structure of the selection statistic. In UVMR, selection thresholds the absolute value of a single *Z*-score, so the conditional distribution of the randomization noise 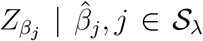 is truncated normal and its moments follow from the Mills ratio. With *p* exposures, the selection statistic (7) becomes a quadratic form whose conditional distribution given the observed data is noncentral 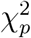 rather than truncated normal, and the Mills ratio is no longer applicable.

In the second step, we construct an initial crude estimate:

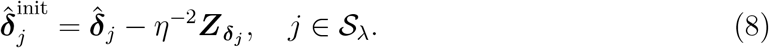

Under this construction, 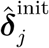 is statistically independent of the selection event 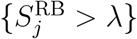 under normality, because 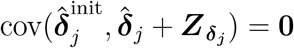 and therefore its submatrix 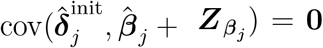. At the same time, we have 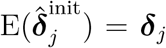. Note that, although the selection event (7) depends only on the exposure estimates, the randomization noise 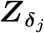 needs to be applied jointly to both the exposure and outcome estimates when the underlying GWAS estimates are correlated. This is a second key distinction from Ma et al.^[5]^, who considered only UVMR with independent GWASs: sample overlap forces the conditional moments to become matrix-valued quantities that couple exposures and outcome GWASs through the cross-covariance 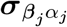. Again, the traditional solution using Mills ratio cannot handle this case, even though *p* = 1.

However, 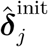 is inefficient because of the added randomization noise. We therefore Rao– Blackwellize it by conditioning on the observed summary statistics and the randomized selection event. Specifically, for each selected variant, define

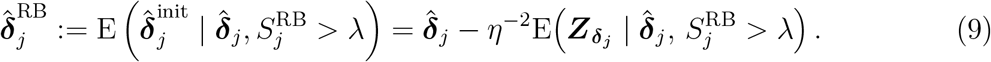

The conditional covariance is defined as 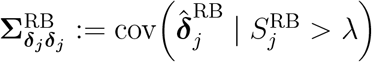 . By the law of total variance,

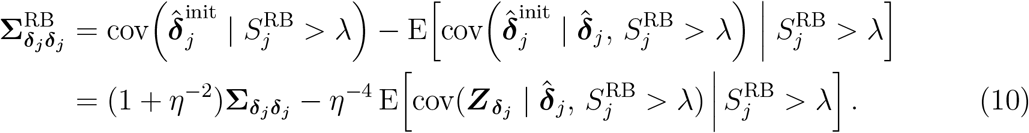

We next derive the conditional mean and covariance of 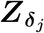 given 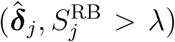, which together yield closed-form expressions for 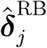 and 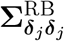. Specifically, conditional on 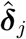, the randomized selection statistic satisfies 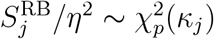, where 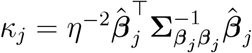, so the required conditional moments can be obtained from the corresponding noncentral *χ*^2^ tail probability. Let 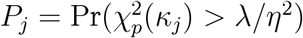 denote this conditional selection probability, and to simplify the conditional-moment formulas, we define the first two derivatives of its log-tail probability as:

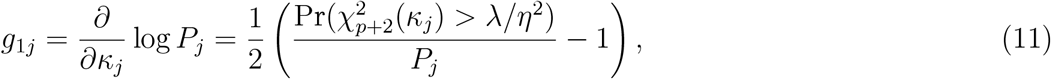

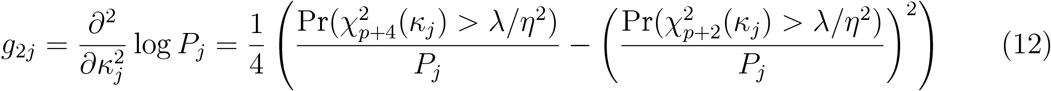

These score functions play analogous roles to the Mills ratio in the univariate case: *g*_1*j*_ governs the direction and magnitude of the conditional mean shift, while *g*_2*j*_ captures the variance contraction due to selection. The conditional mean and covariance matrix of the randomization noise are given by:

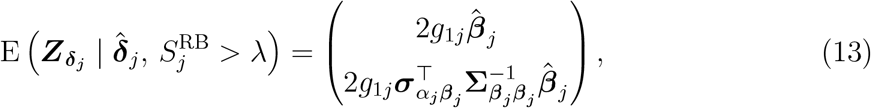

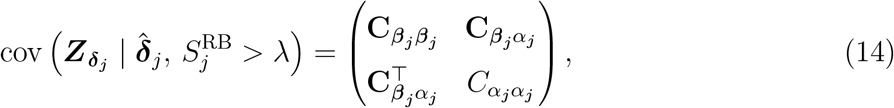

Where

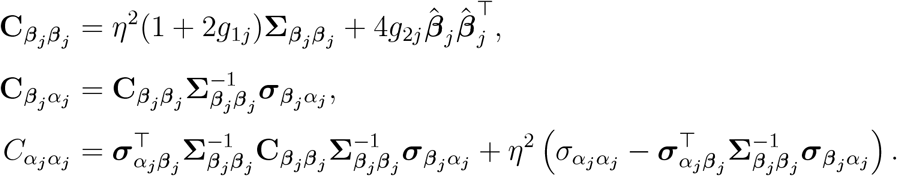

These expressions resolve the inner conditional moments in (10). Nevertheless, the target covariance 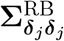 still involves an outer expectation 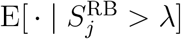 over the marginal law of 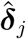, which depends on the unknown true effect ***δ***_*j*_ and is therefore not directly computable. Following the plug-in strategy of Ma et al.^[5]^, we drop this outer expectation and evaluate the inner conditional covariance at the observed 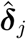, yielding

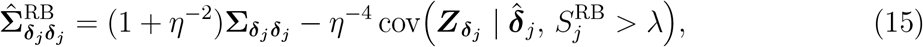

where the conditional covariance takes the closed form (14).

#### Proposition 1

*Suppose* 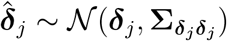 *with* 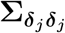 *being positive definite, and* 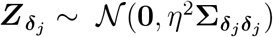 *and is independent of* 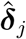. *Then: (i) the initial estimator satisfies* 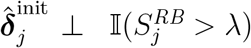; *(ii) the Rao-Blackwellized estimator is conditionally unbiased*, 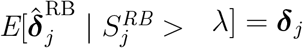; *(iii) the plug-in estimator* 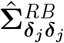 *is conditionally unbiased for* 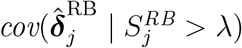. Proposition 1 establishes the validity of the analytical Rao-Blackwellization under the working Gaussian model with a correctly specified covariance 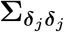. Property (i) is the key construction underlying our procedure: by adding the randomization noise 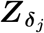 jointly to the exposure and outcome estimates, the initial estimator is shown to be independent of the selection event. This independence implies that the initial estimator remains unbiased after conditioning on selection, and Property (ii) shows that the Rao-Blackwellized estimator preserves this conditional unbiasedness, thereby removing winner’s curse bias. Property (iii) guarantees that the accompanying covariance estimate (15) is conditionally unbiased. Together, these results justify treating the corrected summary statistics 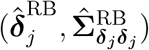 as bias-corrected inputs in downstream MVMR methods. The proof of Proposition 1 is included in Supplementary Material.

We note two practical points regarding the implementation of the multivariable Rao-Blackwellization. First, after re-randomized IV selection based on (7), we will perform LD clumping to ensure that the IVs used in the later MR analysis are approximately independent^[21]^. Second, the multivariable Rao-Blackwellization requires the evaluation of conditional selection probability *P*_*j*_, which is given by the tail of a noncentral *χ*^2^ distribution:

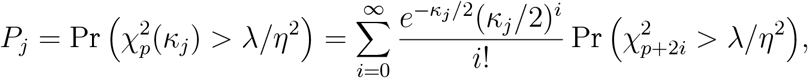

where 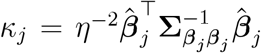 is the noncentrality parameter. When *P*_*j*_ is very small, the event 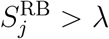 is rare under the observed 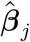, and the conditional moments in (14) become numerically unstable through functions *g*_1*j*_ and *g*_2*j*_. Intuitively, such variants are ones for which selection can be explained by an unusually favorable realization of the randomization noise, making the resulting correction highly sensitive. We therefore exclude variants with *P*_*j*_ *< P*_thres_ (e.g., *P*_thres_ = 0.1 by default) in our implementation, which improves numerical stability and limits the influence of variants whose selection appears to be driven mainly by the randomization noise.

**2.4 Rao-Blackwellized extensions of existing MVMR methods**

The multivariable Rao-Blackwellization provides a flexible plug-and-play module that can be seamlessly integrated into existing MVMR methods. To demonstrate this modularity, we adapt two representative robust MVMR methods: MR-cML^[7,8]^ and MR-BEE^[9]^. We refer to their Rao-Blackwellized modifications as MR-cML-RB and MR-BEE-RB to distinguish them from the original methods that use raw summary statistics.

MR-cML-RB estimates the causal effect vector 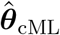 and the pleiotropic effect vector 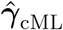 using a constrained likelihood minimization:

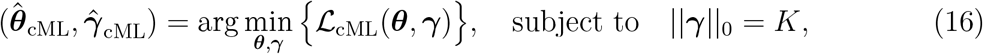

the constraint states that there are *K* IVs having non-zero pleiotropic effects *γ*_*j*_, with the negative log likelihood function

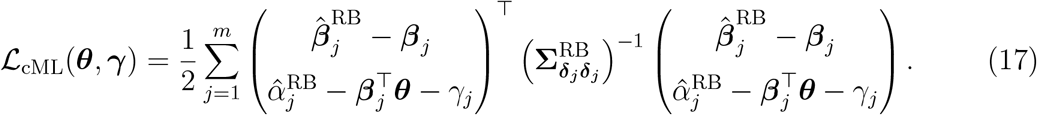

In L_cML_(***θ, γ***), the quantities ***β***_1_, …, ***β***_*m*_ serve as intermediate latent variables, and are jointly updated together with ***θ*** and ***γ*** during minimization. For a given *K*, MR-cML-RB alternates between updating ***θ*** and identifying the *K* IVs whose removal most improves the likelihood until convergence. In practice, MR-cML-RB selects the optimal *K* with the Bayesian information criterion (BIC)^[22]^:

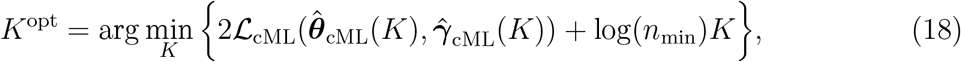

where *n*_min_ is the minimum sample size across GWASs. Following the original method^[8]^, we perform inference via data perturbation. Briefly, we repeatedly generate perturbed versions of the RB-corrected summary statistics from 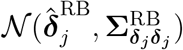, apply MVMR-cML to each perturbed dataset, and obtain the final estimate and covariance matrix as the empirical mean and empirical covariance of the estimates across all perturbed datasets.

MR-BEE-RB lies in its bias-corrected score function, which yields the causal effect estimate 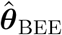 by solving the estimating equation 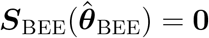, where

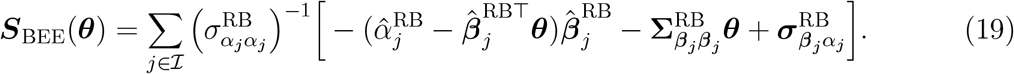

This score function satisfies E(***S***_BEE_(***θ***)) = **0** and hence 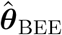 obtained from solving ***S***_BEE_(***θ***_BEE_) = **0** is unbiased. Moreover, I denotes the set of valid IVs (i.e., *γ*_*j*_ = 0) identified through iterative outlier detection^[9,11]^. In implementation, MR-BEE-RB iteratively identifies pleiotropic IVs based on the following hypotheses: for *j* = 1, …, *m*,

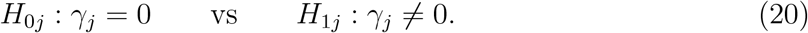

The corresponding test statistic is

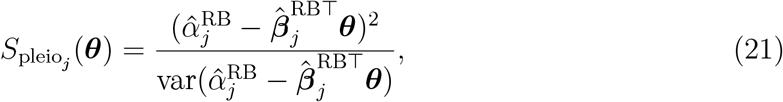

which follows a 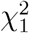 distribution under *H*_0*j*_. Lorincz-Comi et al.^[9]^ introduced robust variance estimation over theoretical variance to capture the effects of underlying balanced pleiotropy, and utilized a false discovery rate (FDR) control method for *p*-value correction^[23]^. Following the original method, we perform statistical inference for MR-BEE-RB via bootstrap^[24]^. Specifically, in each bootstrap replicate, we resample the IVs with replacement from the full set of candidate variants and rerun the entire estimation procedure, including the iterative identification of pleiotropic variants and the estimation of 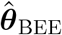. The covariance matrix is then estimated from the empirical covariance of the resulting bootstrap estimates.

## 3 Results

### 3.1 Simulation studies of UVMR

For the UVMR simulations, we considered *M* = 50,000 independent variants. The SNP-exposure effects 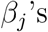 were generated as follows: 1% of variants had nonzero effects generated from N (0, 10^−4^) and then rescaled so that 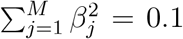, while the remaining variants had *β*_*j*_ = 0. The SNP-outcome effects were set to *α*_*j*_ = *θβ*_*j*_. We considered *N*_*x*_ = *N*_*y*_ = *N* ∈ {30,000, 100,000, 250,000}, and *θ* ∈ {0, 0.1}. 500 replicates were generated for each setting. The observed exposure and outcome GWAS summary statistics were generated as 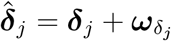, where 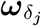 was drawn from a bivariate normal distribution with mean 0 and covariance matrix in (4).

We first examined three scenarios under the UVMR setting: winner’s curse only, winner’s-curse + sample overlap, and winner’s curse + sample overlap + polygenicity. Because IVs are always selected from the same exposure GWAS summary statistics used for estimation, winner’s curse is present in all three scenarios; the latter two settings therefore add sample overlap and polygenic background on top of winner’s curse. In the winner’s-curse-only setting, **R**_***εε***_ = **I**_2_ and **Ω**_*j*_ = **0**. In the sample overlap setting, 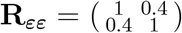 and **Ω**_*j*_ = **0**. In the overlap + polygenicity setting, 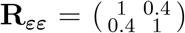 and 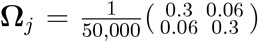 IVs were selected from the exposure GWAS using significance thresholds 5 × 10^−5^ and 5 × 10^−8^, and all methods were evaluated under the two thresholds. We compared MR-BEE-RB and MR-cML-RB with their original counterparts (MR-BEE and MR-cML); MR-APSS and MR-CARE as the most relevant competitors; IVW as the standard UVMR baseline; and the weighted mode estimator, which, alongside MR-APSS, showed well-calibrated type-I error control in the benchmark study of Hu et al. ^[3]^. See Table 1 for a summary of compared MR methods.

As shown in the first set of results in Figure 1A, the winner’s-curse-only setting showed the expected winner’s-curse pattern: methods lacking explicit correction, including MR-BEE, MR-cML, IVW, and the weighted mode, tended to be mildly attenuated, whereas winner’s-curse-aware methods, including MR-BEE-RB, MR-cML-RB, MR-CARE and MR-APSS, largely removed this downward bias. Once the correlation among GWAS estimates was introduced, the separation between methods became much more evident. Across all the sample sizes, MR-BEE-RB and MR-cML-RB remained approximately unbiased and retained close-to-nominal coverage, with MR-APSS showing similarly robust performance. In contrast, MR-cML, MR-BEE, MR-CARE, and IVW became upward biased and undercovered with inflated type-I error in the winner’s-curse + sample-overlap setting, with the distortion becoming even more pronounced after adding polygenic background (Figure 1A-B). Notably, although MR-cML and MR-BEE model sample overlap, they do not correct winner’s curse.

**Figure 1:**
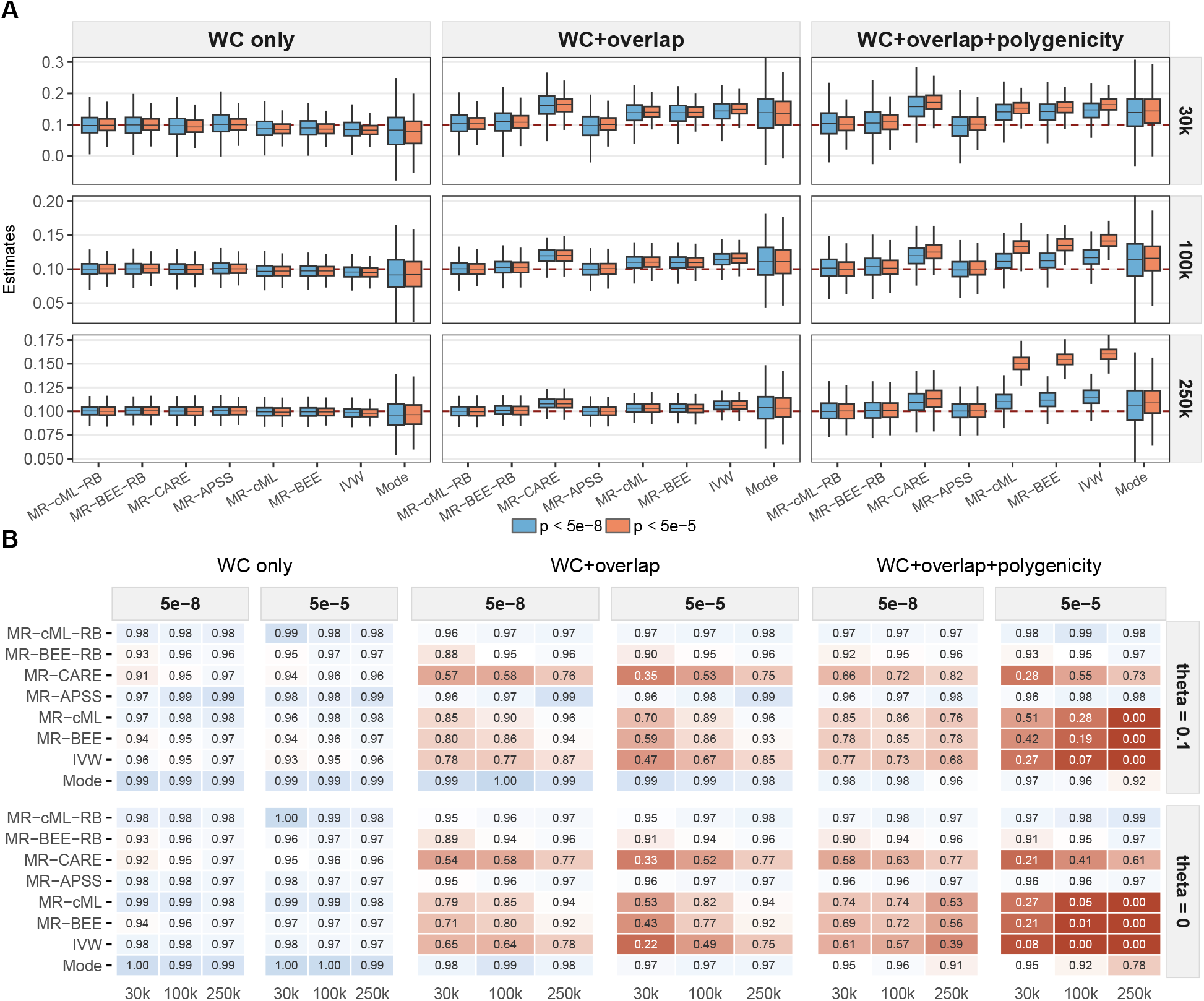
Panel **A** shows the boxplots of estimates at *θ* = 0.1 across 500 replicates, and panel **B** reports the empirical coverage rates across 500 replicates at *θ* = 0.1 and *θ* = 0, under the winner’s-curse-only, winner’s-curse + sample overlap, and winner’s-curse + sample overlap + polygenicity settings.

This pattern shows that accounting for sample overlap alone is not sufficient to remove the bias created by sample overlap and winner’s curse together. Furthermore, MR-CARE also lost accuracy when sample overlap was introduced, because it does not correct the induced bias in the outcome GWAS estimates arising from their correlation with the exposure GWAS estimates, even after correcting winner’s curse in the exposure GWAS estimates. For the weighted mode estimator, upward bias was also observed in the latter two scenarios, but its coverage was relatively better calibrated than that of MR-cML, MR-BEE, MR-CARE, and IVW. Weighted mode also showed slight type-I error inflation in the winner’s-curse + sample-overlap + polygenicity setting. However, the weighted mode estimates were visibly less precise, implying lower power, consistent with the finding in Hu et al. ^[3]^.

In terms of using different *p*-value thresholds for IV selection, moving from 5 × 10 ^−8^ to 5 × 10 ^−5^ had relatively little effect on MR-BEE-RB, MR-cML-RB, and MR-APSS, and had a modest effect on the weighted mode in Figure 1A. However, Figure 1 B shows that the weighted mode still lost some calibration under the more liberal threshold in the winner’s-curse + sample-overlap + polygenicity setting. The largest deterioration was seen for MR-cML, MR-BEE, MR-CARE, and IVW, whose bias and undercoverage increased substantially once a polygenic background was present, especially at larger *N* . This is because, as *N* increased, the looser threshold included many more SNPs whose apparent associations are likely driven by polygenic background and random noise rather than the sparse causal signal. In the sample overlap + polygenicity setting, the proportion of selected null/background SNPs increased from about 12%, 26%, and 64% at *N* = 30,000, *N* = 100,000, and *N* = 250,000, respectively, under the 5 × 10 ^−5^ threshold.

We also conducted an additional UV simulation with 10% invalid IVs exerting both uncorrelated and correlated pleiotropy, and the full results are reported in the Supplementary Materials. Briefly, in the winner’s-curse-only setting, MR-BEE-RB, MR-cML-RB, and MR-CARE remained unbiased, whereas MR-APSS became biased. The possible reason is that MR-APSS’s foreground model assumes that the direct effect on the outcome is independent of the instrument strength, i.e., it does not accommodate correlated pleiotropy in its foreground model. When the sample overlap and polygenicity were introduced, only MR-BEE-RB and MR-cML-RB remained unbiased, which further supports the robustness of the proposed methods when winner’s curse, invalid IVs, sample overlap and polygenic background are all present.

### 3.2 Simulation studies of MVMR

Next, we considered 3 exposures to demonstrate the proposed procedure in the MVMR setting. The true causal effect vector was set to ***θ*** = (0.2, 0.1, 0) ^⊤^. We generated sparse genetic effects ***β***_*j*_ for 1% of variants out of *M* = 50 000 under a mixed architecture allowing a variant to affect one, two, or all three exposures. On the active set, the nonzero entries of ***β***_*j*_ were drawn from a multivariable normal distribution with correlation matrix 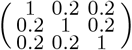, and were then rescaled so that 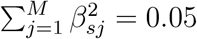 for each exposure. The outcome effects were set to *α*_*j*_ = *β*_1*j*_*θ*_1_ + *β*_2*j*_*θ*_2_ + *β*_3*j*_*θ*_3_. For the main-text figure, we focused on the winner’s-curse-only setting and the winner’s-curse + overlap + polygenicity setting. Additional results under the winner’s-curse + sample overlap scenario and invalid IV settings are reported in the Supplementary Material. As in the UVMR simulations, winner’s curse remains present in both settings because IVs are selected from the same simulated exposure GWAS summary statistics. For the winner’s-curse-only setting, **R**_***εε***_ = **I**_4_ and **Ω**_*j*_ = **0**. In the winner’s-curse + sample overlap + polygenicity setting, we set

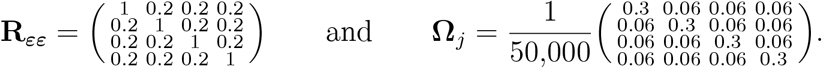

IVs were selected using the joint association statistic across the three exposure GWASs. As in the univariable setting, we examined thresholds 5 × 10 ^−5^ and 5 × 10 ^−8^, and three sample sizes of *N* ∈ {30,000, 100,000, 250,000}. For each simulation set-up, we ran 500 replications and compared MR-BEE-RB, MR-cML-RB, and existing MVMR methods including MR-BEE, MR-cML, MR-IVW and MR-Median.

Figure 2A shows that, in the winner’s-curse-only setting with no correlation among GWAS estimates, methods without Rao-Blackwellization showed mild attenuation for the nonzero exposure *X*_1_ (and *X*_2_, Supplemental Figure S4), especially at relatively small sample sizes. This downward bias was most evident for IVW and the weighted median at *N* = 30,000, and gradually diminished as *N* increased to 100,000 and 250,000. For the null exposure *X*_3_, all methods remained centered close to 0. The corresponding coverage results in Figure 2C show that coverage under winner’s curse alone was generally close to nominal.

**Figure 2:**
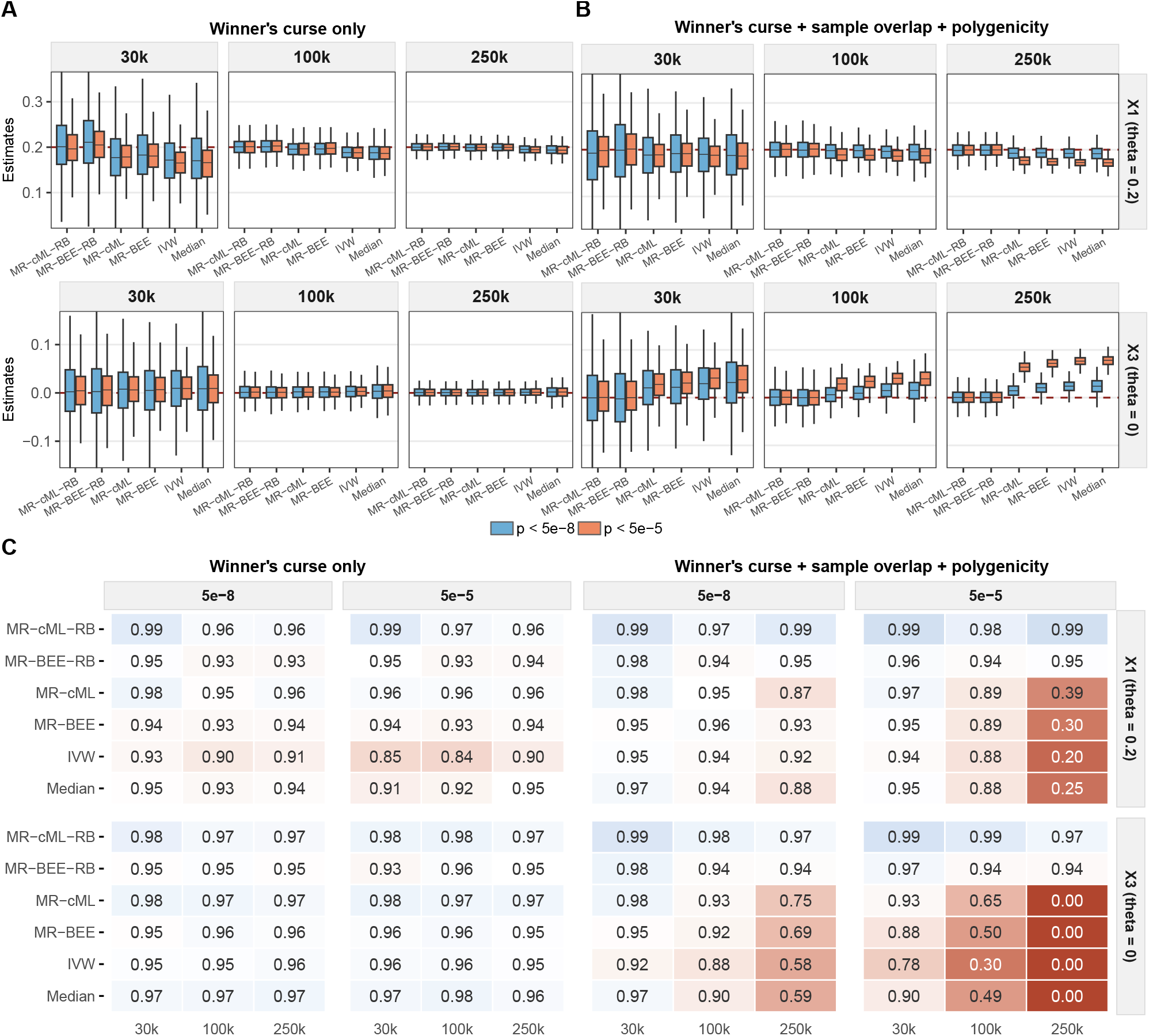
Panels **A-B** show the boxplots of estimates for *X*_1_ and *X*_3_ across 500 replicates under (**A**) winner’s-curse-only and (**B**) winner’s-curse + sample overlap + polygenicity, respectively. Panel **C** reports the empirical coverage rates across 500 replicates at *θ*_1_ = 0.2 and *θ*_3_ = 0, under the two settings. Results for *X*_2_ are provided in the Supplementary Material.

Under winner’s-curse + sample overlap + polygenicity (Figure 2B), the difference between the Rao-Blackwellized and uncorrected methods became much more pronounced. MR-BEE-RB and MR-cML-RB remained almost unbiased for all exposures (see results for *X*_2_ in Supplemental Figure S7) across all sample sizes and both IV-selection thresholds. In contrast, the non-RB methods became increasingly biased as *N* increased, and the bias was worse under the liberal threshold 5 × 10 ^−5^. For the nonzero exposure *X*_1_, the non-RB methods showed clear downward bias, whereas for the null exposure *X*_3_ they showed substantial upward bias. Substantial undercoverage for non-RB methods was also observed, most clearly under the liberal threshold (5 × 10 ^−5^) and with larger sample sizes, while MR-BEE-RB and MR-cML-RB remained close to the nominal 95% level (Figure 2C). Overall, the MV results again highlighted the robustness of MR-BEE-RB and MR-cML-RB to the winner’s curse and polygenic background.

Finally, we compared the two common IV selection strategies used in MVMR, the joint-selection strategy based on a multivariable association test and the exposure-specific screening strategy followed by taking the union across exposures (termed the UV-then-union strategy), as described in Section 2.2. As shown in Supplemental Figure S8, the two strategies resulted in similar results under the stringent threshold 5 × 10 ^−8^. Under the liberal threshold 5 × 10 ^−5^, the UV-then-union strategy was generally more biased than the joint-selection strategy, likely because it included more weak or polygenicity-driven variants.

### 3.3 UVMR benchmark in real data positive and negative control analyses

We evaluated the UVMR methods using similar positive and negative control frameworks as Hu et al. ^[3]^. The positive control (i.e., same-trait analysis) assesses whether a method can recover a causal effect of 1, whereas the negative control analysis assesses whether a method remains well calibrated when no causal effect is expected. We compared MR-cML-RB, MR-BEE-RB, MR-cML, MR-BEE, and the benchmark competitors MR-APSS, MR-CARE, IVW, and the weighted mode estimator in the analyses.

For the positive control analysis, we considered 33 quantitative traits from the UK Biobank, where each trait was used as both exposure and outcome in UVMR. In this setting, the true causal effect is expected to be 1. We considered two sample-overlap settings when generating the exposure and outcome GWAS summary statistics: no sample overlap and 50% sample overlap. IVs were selected using both the conventional threshold of 5 × 10 ^−8^ and the liberal threshold of 5 × 10 ^−5^. For readability, we did not report the 5 × 10 ^−5^ results for non-winner’s-curse-aware methods in the main-text figures. Instead, these results were presented in Supplemental Figure S9. In addition, we computed the LD scores *l*_*j*_ using the LD reference panel of Zheng et al. ^[25]^, which comprises *M* ≈ 7.35 million SNPs. Details of the generation and harmonization of GWAS summary statistics are provided in the Supplementary Material.

Figure 3A presents the estimated effects and nominal 95% confidence intervals in the no-sample-overlap setting. In this setting, the uncorrected MR-BEE, MR-cML, and IVW methods showed the expected downward bias from winner’s curse, with many estimates falling below 1. The weighted mode estimator was generally less biased than MR-BEE, MR-cML, and IVW, but it was also less precise, as reflected by its widest intervals among the methods. In contrast, MR-BEE-RB and MR-cML-RB achieved the best overall coverage among the methods. MR-APSS and MR-CARE also reduced the winner’s curse bias with comparable coverage rates, but their estimates were more variable with wider confidence intervals than both MR-BEE-RB and MR-cML-RB. Supplemental Figure S9C further shows that MR-BEE-RB and MR-cML-RB were computationally more efficient than MR-APSS and MR-CARE.

**Figure 3:**
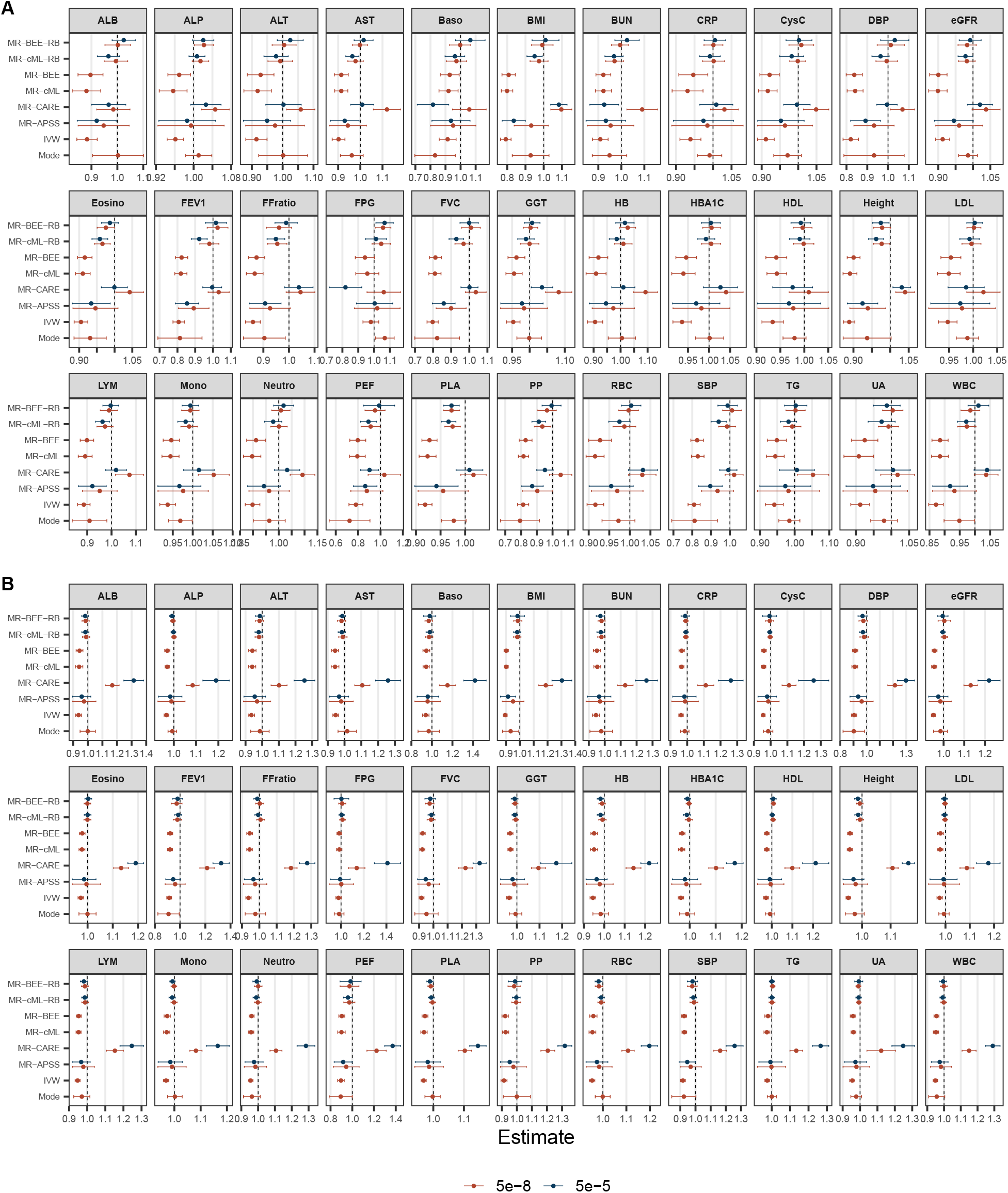
Causal effect estimates and computational runtime from the same-trait analysis. **(A)** Causal effect estimates and nominal 95% confidence intervals in the 0% overlap setting **(B)** Corresponding results in the 50% sample overlap setting. In both panels, results are shown under IV selection thresholds of 5 ×10 ^−5^ (blue) and 5 ×10 ^−8^ (red). The dashed horizontal line indicates the true causal effect of 1.

Across IV-selection thresholds, MR-BEE-RB showed the most stable overall performance, whereas MR-cML-RB and MR-APSS appeared better at a more stringent threshold of 5 × 10 ^−8^, and MR-CARE performed well at 5 × 10 ^−5^. Supplemental Figure S9A further shows that, under the liberal threshold of 5 × 10 ^−5^, MR-BEE, MR-cML, IVW, and the weighted mode became much more downward biased, consistent with stronger winner’s-curse distortion when weaker instruments are included.

Figure 3B presents the results in the 50% overlap setting. Across both thresholds, MR-cML-RB and MR-BEE-RB remained centered closest to 1 and showed the best overall calibration, while MR-APSS performed similarly in terms of bias correction but again yielded visibly wider confidence intervals. The weighted mode estimator also yielded comparatively good coverage, although its estimates were less precise. In contrast, MR-CARE, which performed reasonably well in the no-overlap setting, became clearly upward biased under sample overlap, which is expected because it does not correct the induced bias in the outcome GWAS estimates arising from their correlation with the exposure GWAS estimates. The uncorrected MR-cML, MR-BEE, and IVW methods remained downward biased, and Supplemental Figure S9 further shows that this downward bias became more pronounced under the liberal threshold. Moreover, the apparent absence of bias for weighted mode in 50% overlap case may reflect a cancellation artifact: weak-instrument bias (downward) and sample-overlap bias (upward) offset each other near this point, with sample-overlap bias dominating as the overlap increases ^[9]^.

We next considered a negative control analysis. Specifically, we paired the same 33 quantitative traits used in the previous section with three negative-control outcomes in UK Biobank, including natural hair color before greying (black), natural hair color before greying (red), and liking for coriander, which yielded a total of 99 UVMR analyses. These outcomes are primarily influenced by germline variation in pigmentation genes (*MC1R, HERC2* /*OCA2* ) and olfactory receptor genes (*OR6A2* ) ^[26,27]^. They are therefore unlikely to be downstream consequences of the 33 exposure traits considered here and were used as negative-control outcomes. Details of GWAS summary statistics generation and harmonization are provided in the Supplementary Material.

Figure 4 summarizes the method-specific QQ plots of −log_10_(*p*) values from the negative control analyses under IV selection thresholds of 5 × 10 ^−5^ (blue) and 5 × 10 ^−8^ (red). Overall, MR-cML-RB achieved the best calibration across both thresholds. MR-BEE-RB showed some inflation, with more under 5 × 10 ^−8^ than under 5 × 10 ^−5^. The weighted mode estimator was well calibrated with only slight tail inflation. MR-APSS remained reasonably well calibrated under 5 × 10 ^−8^, but showed some inflation under 5 × 10 ^−5^. In contrast, MR-CARE, MR-cML, MR-BEE and IVW exhibited substantial inflation, and the inflation was more pronounced under the liberal threshold of 5 × 10 ^−5^. Taken together, the two UVMR benchmark analyses suggest that, although the liberal threshold 5 × 10 ^−5^ may still be reasonable to use with the winner’s-curse-aware procedures, the stringent threshold 5 × 10 ^−8^ was more stable across methods and overlap settings than the liberal threshold, and therefore appears to be the safer default for routine analyses.

**Figure 4:**
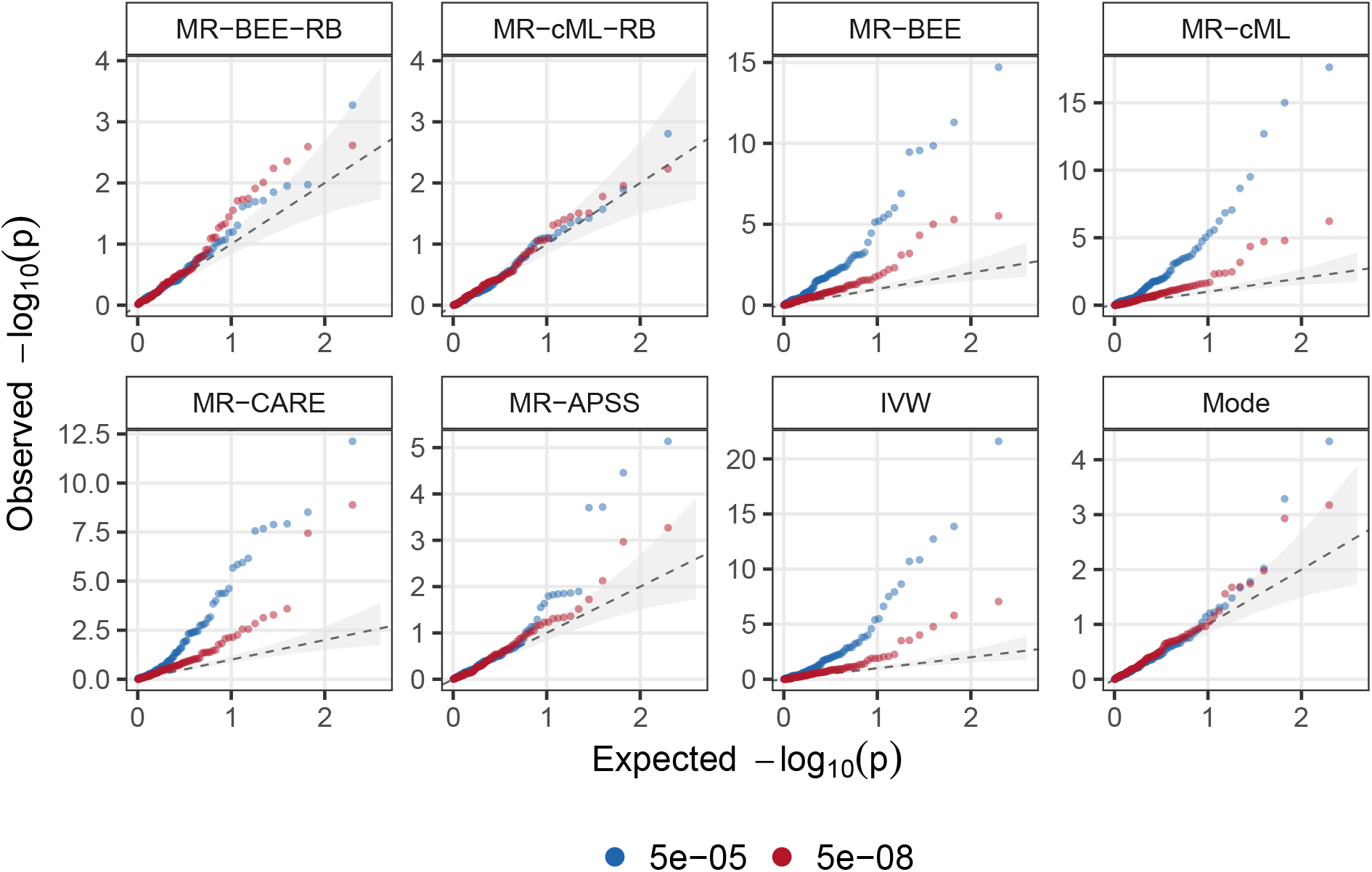
QQ plots of − log_10_(*p*) values in negative control outcome analysis. The shaded regions represent the 95% confidence interval. Results are shown under IV selection thresholds of 5 × 10 ^−5^ (blue) and 5 × 10 ^−8^ (red).

### 3.4 MVMR analysis of risk factors on CAD

The causal role of HDL cholesterol on CAD remains one of the most debated questions in cardiovascular genetics. Despite strong observational associations, clinical trials of pharmacological agents designed specifically to raise HDL cholesterol have repeatedly failed to reduce CAD risk ^[28]^. Even in the case of *CETP* inhibitors, which produce dramatic HDL cholesterol elevation alongside moderate LDL cholesterol reduction, the cardiovascular benefit of anacetrapib is now widely attributed to its LDL cholesterol-lowering effect rather than to HDL cholesterol elevation per se ^[29,30]^. Early UVMR analyses were similarly null ^[31]^, yet more recent studies leveraging the large meta-analysis ^[32]^ have reported significant protective estimates for HDL cholesterol on CAD ^[9,10,33]^. We hypothesize that this discrepancy is driven by the uncontrolled winner’s curse and polygenic background, which inflate HDL cholesterol effect estimates. To investigate this, we performed MVMR analysis using MR-BEE-RB and MR-cML-RB for 10 cardiometabolic exposures on CAD, including body mass index (BMI), smoking initiation (SMK), drinks per week (DRNK), diastolic blood pressure (DBP), systolic blood pressure (SBP), HDL cholesterol, LDL cholesterol, TG, T2D, and urate (UA). Details of data generation and harmonization are presented in the Supplementary Material.

Figure 5 presents the MVMR estimates of 10 cardiometabolic risk factors on CAD under both the traditional and Rao-Blackwellization-corrected analyses. The most notable finding concerns HDL cholesterol, whose estimated protective effect on CAD was substantially attenuated after correction: the traditional estimates were −0.130 (SE = 0.035) and −0.110 (SE = 0.019) for MR-cML and MR-BEE, respectively, and were both statistically significant, whereas the corrected estimates reduced to −0.075 (SE = 0.041) and −0.077 (SE = 0.042), and were no longer significant. Our result suggested that the significant protective effect of HDL cholesterol on CAD in previous studies could be attributable to winner’s curse and background polygenicity rather than a true causal effect. In contrast, the estimated effect of TG on CAD was strengthened after correction, from non-significant traditional estimates of 0.029 (SE = 0.033) and 0.038 (SE = 0.019) to significant estimates of 0.073 (SE = 0.034) and 0.064 (SE = 0.031) for MR-cML and MR-BEE, respectively, suggesting that the traditional approach underestimates the causal effect of TG. For SBP and LDL cholesterol, the estimates were consistent across both methods and both analyses, providing robust evidence of their causal effects on CAD. Finally, for SMK, the traditional estimates differed substantially between MR-cML (0.395, SE = 0.083) and MR-BEE (0.224, SE = 0.032), whereas the corrected estimates converged to 0.279 and 0.247, respectively, suggesting that the Rao–Blackwellization correction stabilizes the estimation in the presence of winner’s curse and background bias.

**Figure 5:**
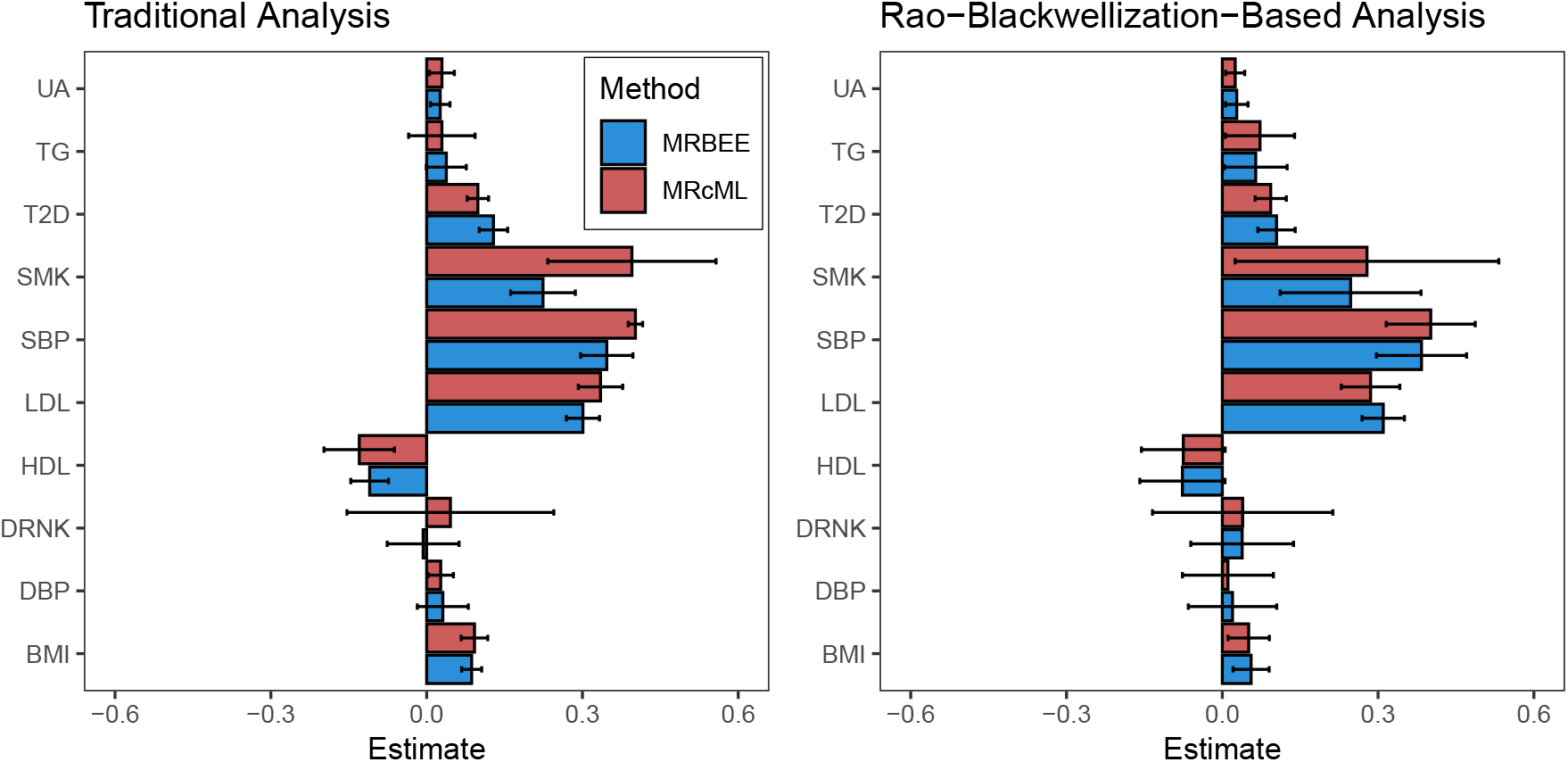
Estimated causal effects and 95% confidence intervals of 10 cardiometabolic risk factors on CAD. Left: Results from the traditional analysis based on 3,198 independent IVs selected without Rao-Blackwellization correction. Right: Results after applying Rao-Blackwellization correction based on 1,217 independent IVs. Each panel displays point estimates with 95% confidence intervals for MVMR analyses using MR-BEE (blue) and MR-cML (red).

## 4 Discussion

The main methodological contribution of this paper is an analytical, closed-form multi-variable Rao-Blackwellization that corrects winner’s curse while accounting for polygenic background and sample structure in GWAS summary statistics. Rather than introducing a new method-specific MR estimator, the proposed framework produces bias-corrected summary statistics and covariance estimates with explicit theoretical guarantees, which can be used as a plug-in correction layer in both UVMR and MVMR analyses. When combined with downstream robust estimators such as MR-cML and MR-BEE, the proposed corrections improve robustness to winner’s curse, sample overlap, and polygenic background without requiring fundamental changes to the core estimation procedures. Protection against weak-instrument bias and horizontal pleiotropy is then inherited from the chosen downstream MR method.

Our simulations systematically characterize how winner’s curse, sample overlap, and polygenicity distort existing MR methods across the UVMR and MVMR settings and IV-selection thresholds. Winner’s curse alone generally produced mild attenuation when the causal effect size was small, whereas larger biases and poorer coverage emerged after sample overlap and polygenic background were introduced. Polygenicity was especially influential under the liberal 5 × 10 ^−5^ threshold: many weak-signal and null variants enter the instrument set, with contamination worsening as the sample size grows, explaining why larger GWAS do not necessarily guarantee more reliable causal inference. Our work demonstrates that MR-BEE-RB and MR-cML-RB are well calibrated and unbiased across the examined settings. MR-APSS also performed comparatively well in most UVMR settings, and the weighted mode often retained reasonable calibration but yielded slightly biased and less precise estimates, broadly consistent with the benchmark findings of Hu et al. ^[3]^. These findings suggest that the 5 × 10 ^−8^ threshold is still a safer default in practice, which sacrifices only a little efficiency relative to the 5 × 10 ^−5^ threshold in ideal settings but is more robust when the assumptions are violated.

The UVMR positive and negative control analyses showed similar overall patterns in empirical data. MR-BEE-RB, MR-cML-RB, and MR-APSS were generally among the better-calibrated methods, although MR-APSS tended to produce wider confidence intervals. The weighted mode was well calibrated in the negative control analysis, but still exhibited winner’s curse bias in the same-trait analysis. MR-CARE performed well in removing winner’s curse under the no sample overlap scenario in the same-trait analysis, but failed to maintain coverage under 50% sample overlap. Moreover, we independently performed the UVMR benchmark analyses twice on different computing platforms and obtained consistent results (Supplemental Figures S10-S11), providing evidence that the randomized selection step was stable for these analyses. We also explored an additional winner’s-curse-aware method, MRlap ^[20]^, in the benchmark studies. However, using its recommended pipeline, we encountered practical difficulties, including unresolved implementation issues and prohibitively long runtime. We therefore did not include MRlap in the benchmark comparison. In the MVMR analysis of ten cardiometabolic risk factors on CAD, the estimated protective effect of HDL cholesterol was substantially attenuated after correction and was no longer statistically significant with either MR-cML-RB or MR-BEE-RB, suggesting that this effect is more likely driven by uncontrolled winner’s curse and background noise. The correction also revealed and stabilized causal signals for other risk factors that were obscured or unstable under the traditional analysis.

Several future directions merit discussion. First, the current framework treats the covariance components estimated by LDSC as fixed plug-in quantities. Although Hu et al. ^[4]^ has evaluated the influence of this estimation uncertainty using a block-wise jackknife over genome-wide SNPs and found similar inference with and without the resulting standard-error adjustment in the UVMR simulation settings considered, future work needs to investigate how to propagate the uncertainty in the LDSC estimates through Rao-Blackwellization and downstream causal inference. Second, in practice, particularly in MVMR analyses, joint selection may retain IVs that are effectively weak for a specific exposure, where the estimation error 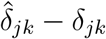 is large relative to the true effect *δ*_*jk*_ for the *k*th exposure. This typically arises when the GWAS of the *k*th exposure has limited power due to small sample size or low trait heritability. Under such weak instruments, MR-BEE and MR-cML can become unstable, producing estimates with inflated effect sizes accompanied by even larger SEs. To solve this issue, we apply an empirical procedure from our previous work ^[34]^ to limit the influence of these IVs (Supplementary Material). Third, the present work focuses exclusively on the application of Rao-Blackwellization in the standard summary-data-based MR setting. A natural extension is to adapt this framework to other MR paradigms, including *cis*-MR ^[35,36,37,38]^, multi-ancestry MR ^[39,40]^, and network MR ^[15,34,41,42]^, where analogous polygenicity- and selection-induced biases may arise. Lastly, during the preparation of this manuscript, Forde et al. ^[43]^ proposed MR-SimSS that uses repeated simulated sample splitting in UVMR to address biases arising from instrument selection, weak instruments, and participant overlap. Conceptually, the two methods are related: both aim to reduce bias caused by IV selection, but MR-SimSS relies on repeated stochastic reconstruction of independent pseudo-samples, while our method directly yields corrected GWAS association estimates and the covariance matrices. Extending this alternative correction strategy to MVMR and settings with a polygenic background is an important direction for future research.

## Data Availability

All GWAS summary statistics used in this study are publicly available, with their sources listed in Table S1 and Table S2. The individual-level data from the UK Biobank used for analyses are available under Application ID: 81097.

## Acknowledgments

This work was supported by grants HG011052 and HG011052-03S1 from the National Human Genome Research Institute (NHGRI), USA (to X.Z.); grant HL086694 from the National Heart, Lung, and Blood Institute (NHLBI), USA; and the Florida State University CTSA Provost award (to Z.L.).

## Data and Code Availability

All GWAS summary statistics used in this study are publicly available, with their sources listed in Table S1 and Table S2. The individual-level data from the UK Biobank used for analyses are available under Application ID: 81097. The R package RBCorrection, which implements the method, is available at https://github.com/harryyiheyang/RBCorrection.

## Notes

### Competing Interest Statement

The authors have declared no competing interest.

