## Supplementary Materials for "Recalibrating Mendelian randomization under winner’s curse, sample structure and polygenicity"

Yihe Yang†
Department of Population and Quantitative Health Sciences,
Case Western Reserve University School of Medicine
Zhaotong Lin†,*
Department of Statistics, Florida State University
Haoran Xue
Department of Biostatistics, City University of Hong Kong
Xiaofeng Zhu*
Department of Population and Quantitative Health Sciences,
Case Western Reserve University School of Medicine

†These authors contributed equally to this work.

**S4 Implementation details and tuning parameters. 22**

### Supplemental Figures

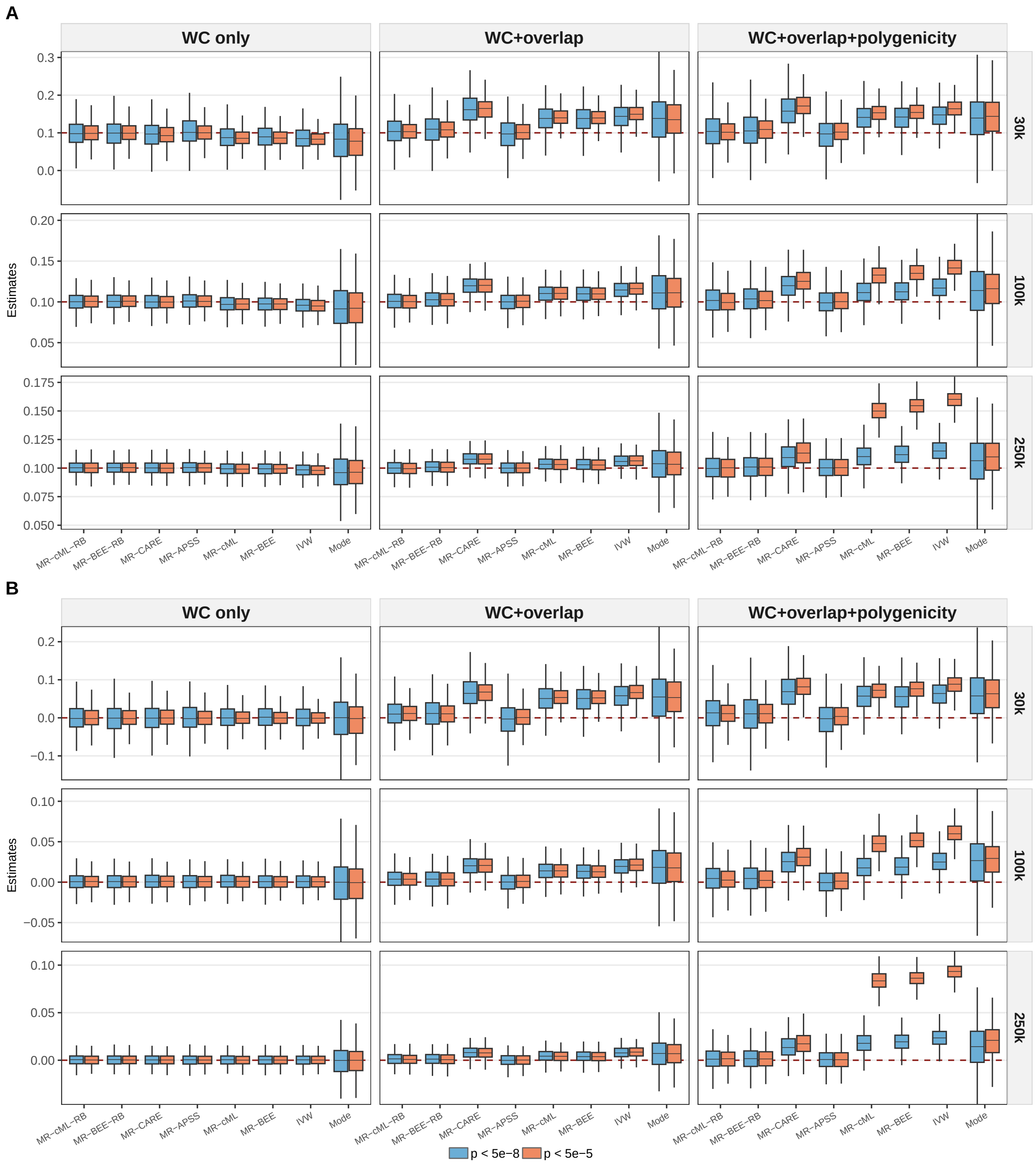

Figure S1: Boxplots of causal effect estimate in the benchmark UV setting with no invalid IV. Panels **A** and **B** corresponded to $\theta = 0$ and $\theta = 0.1$, respectively. Within each panel, results were shown across the no-background, overlap-only, and overlap + polygenic settings, for IV selection thresholds $5 \times 10^{-8}$ and $5 \times 10^{-5}$, and sample sizes $N = 30{,}000$, $100{,}000$, and $250{,}000$.

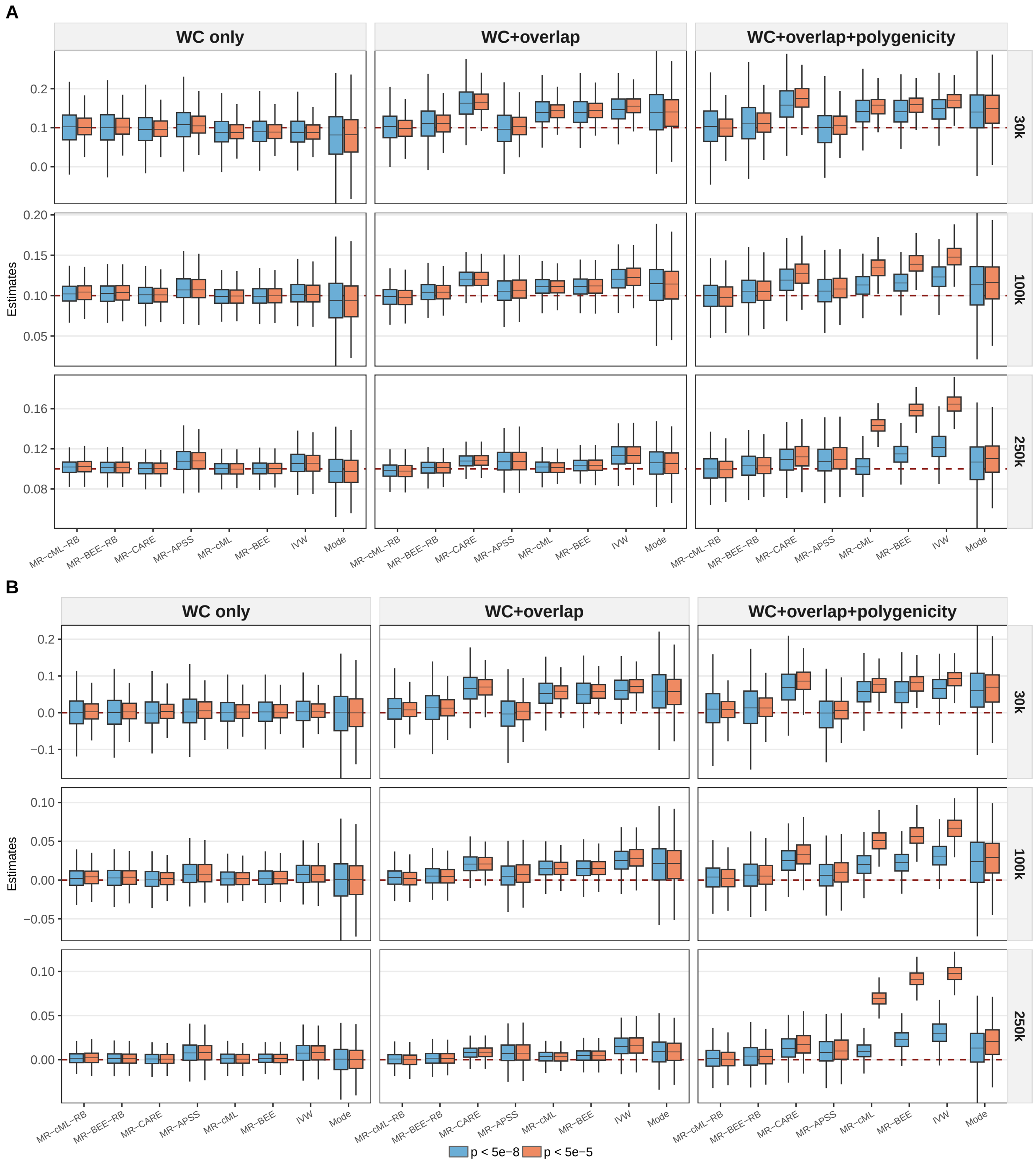

Figure S2: Boxplots of causal effect estimate in the additional UV simulation with 10% invalid IVs. Panels **A** and **B** corresponded to $\theta = 0$ and $\theta = 0.1$, respectively. Within each panel, results were shown across the no-background, overlap-only, and overlap + polygenic settings, for IV selection thresholds $5 \times 10^{-8}$ and $5 \times 10^{-5}$, and sample sizes $N = 30{,}000$, 100,000, and 250,000.

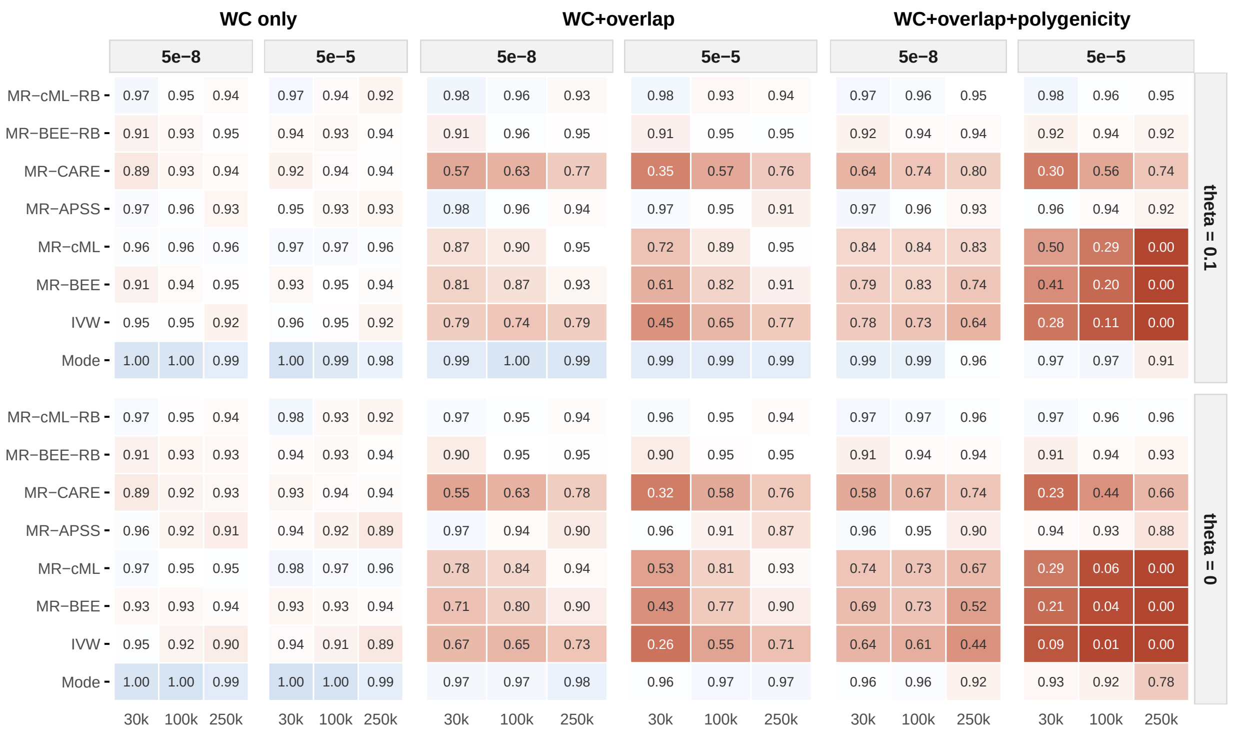

Figure S3: Empirical coverage rates across 500 replicates in the additional UV simulation with 10% invalid IVs. The two rows corresponded to $\theta = 0.1$ and $\theta = 0$, and the three blocks corresponded to the winner's-curse-only, winner's curse + sample-overlap, and winner's curse + overlap + polygenic settings. Within each block, the two subcolumns corresponded to IV selection thresholds $5 \times 10^{-8}$ and $5 \times 10^{-5}$, and columns corresponded to sample sizes $N = 30{,}000$, 100,000, and 250,000.

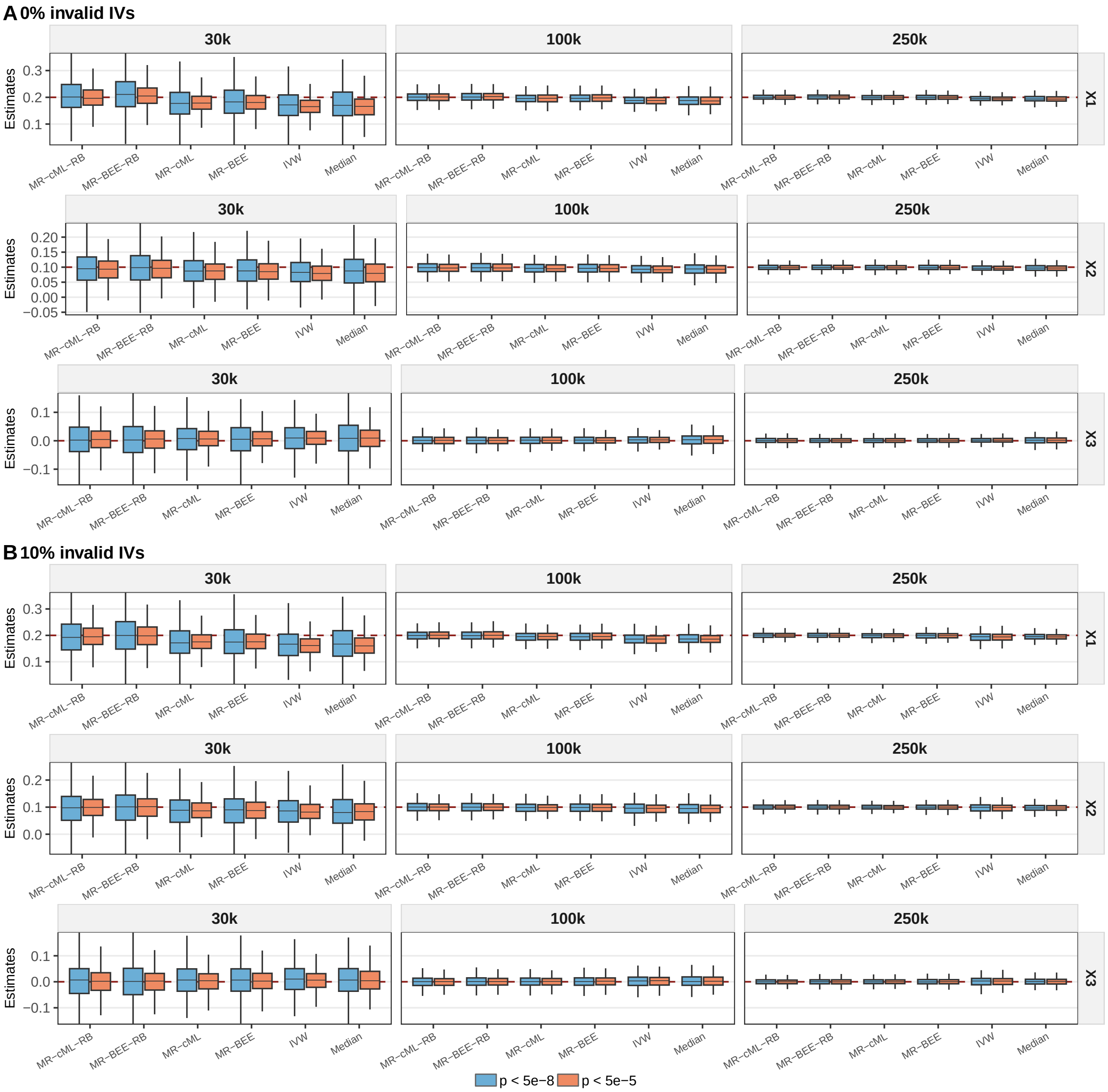

Figure S4: Boxplots of MVMR estimates in the winner's-curse-only setting. Panels **A** and **B** corresponded to the 0% and 10% invalid-IV settings, respectively. Within each panel, rows corresponded to $X_1$ ($\theta_1 = 0.2$), $X_2$ ($\theta_2 = 0.1$), and $X_3$ ($\theta_3 = 0$), columns corresponded to sample sizes $N = 30{,}000$, $100{,}000$, and $250{,}000$, and results were shown for IV selection thresholds $5 \times 10^{-8}$ and $5 \times 10^{-5}$.

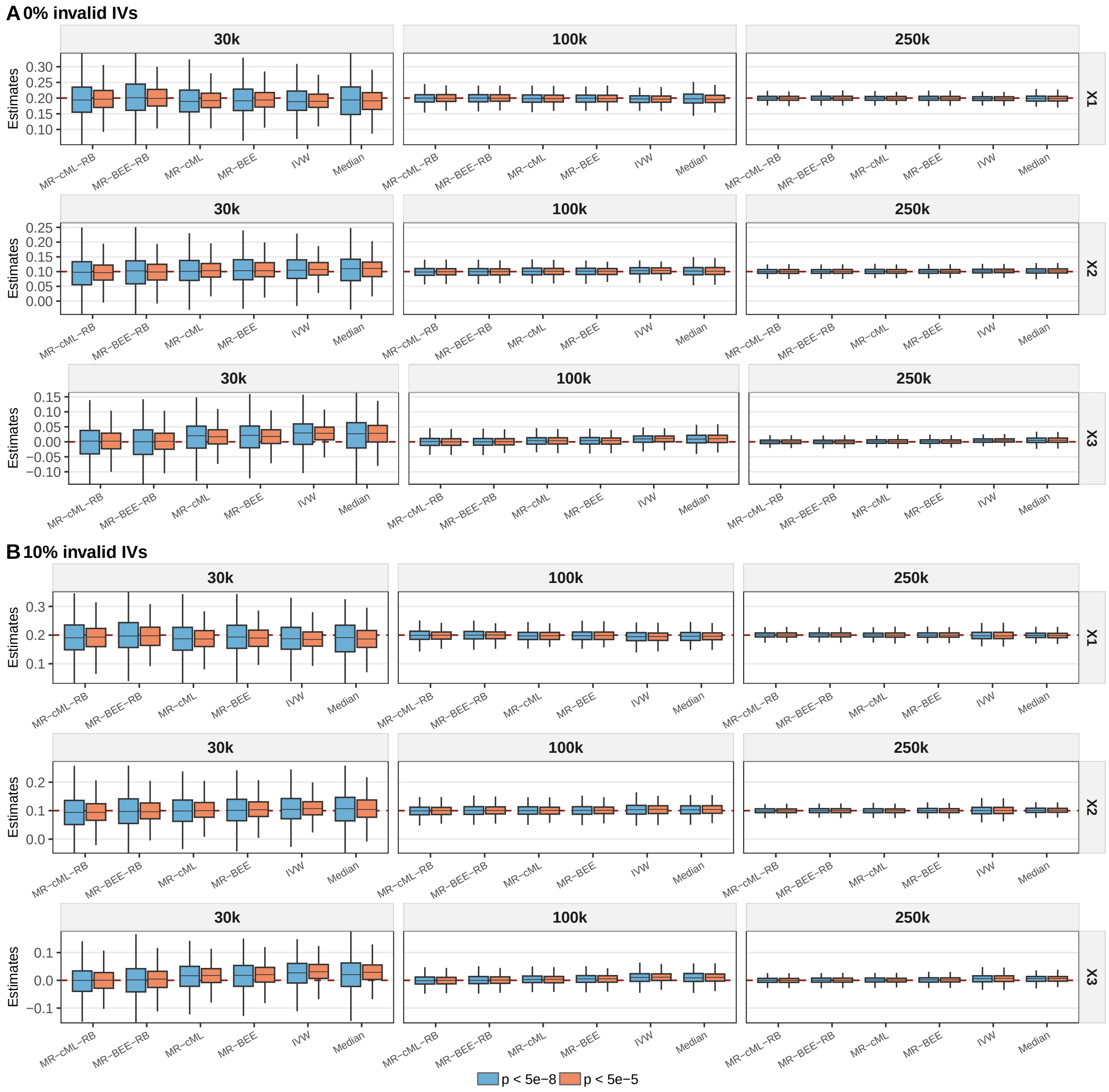

Figure S5: Boxplots of MVMR estimates in the winner's curse + sample-overlap setting. Panels **A** and **B** correspond to the 0% and 10% invalid-IV settings, respectively. Within each panel, rows correspond to $X_1$, $X_2$, and $X_3$, columns correspond to sample sizes $N = 30{,}000$, 100,000, and 250,000, and results are shown for IV selection thresholds $5 \times 10^{-8}$ and $5 \times 10^{-5}$.

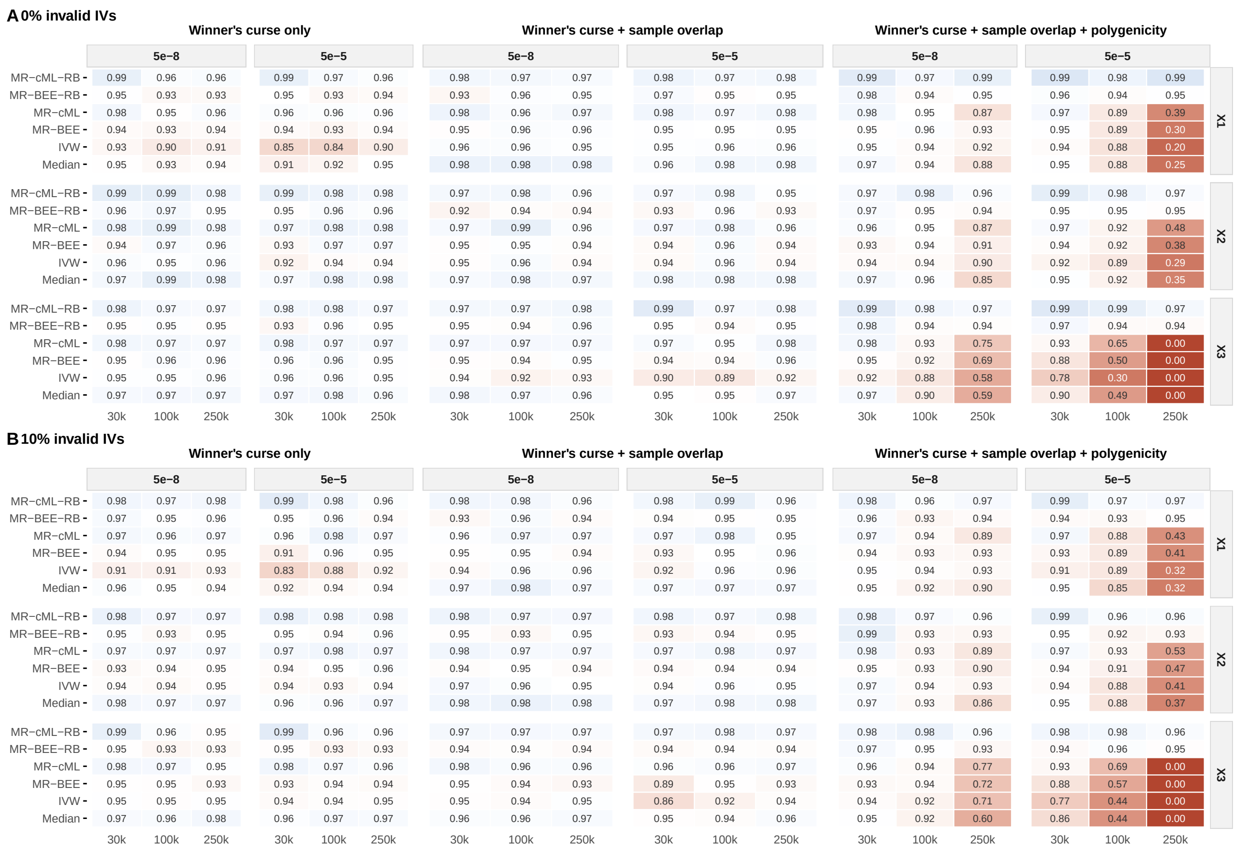

Figure S6: Empirical coverage rates across all MVMR scenarios. Panels **A** and **B** correspond to the 0% and 10% invalid-IV settings, respectively. From left to right correspond to the winner's-curse-only, winner's curse + sample-overlap, and winner's curse + sample-overlap + polygenicity settings.

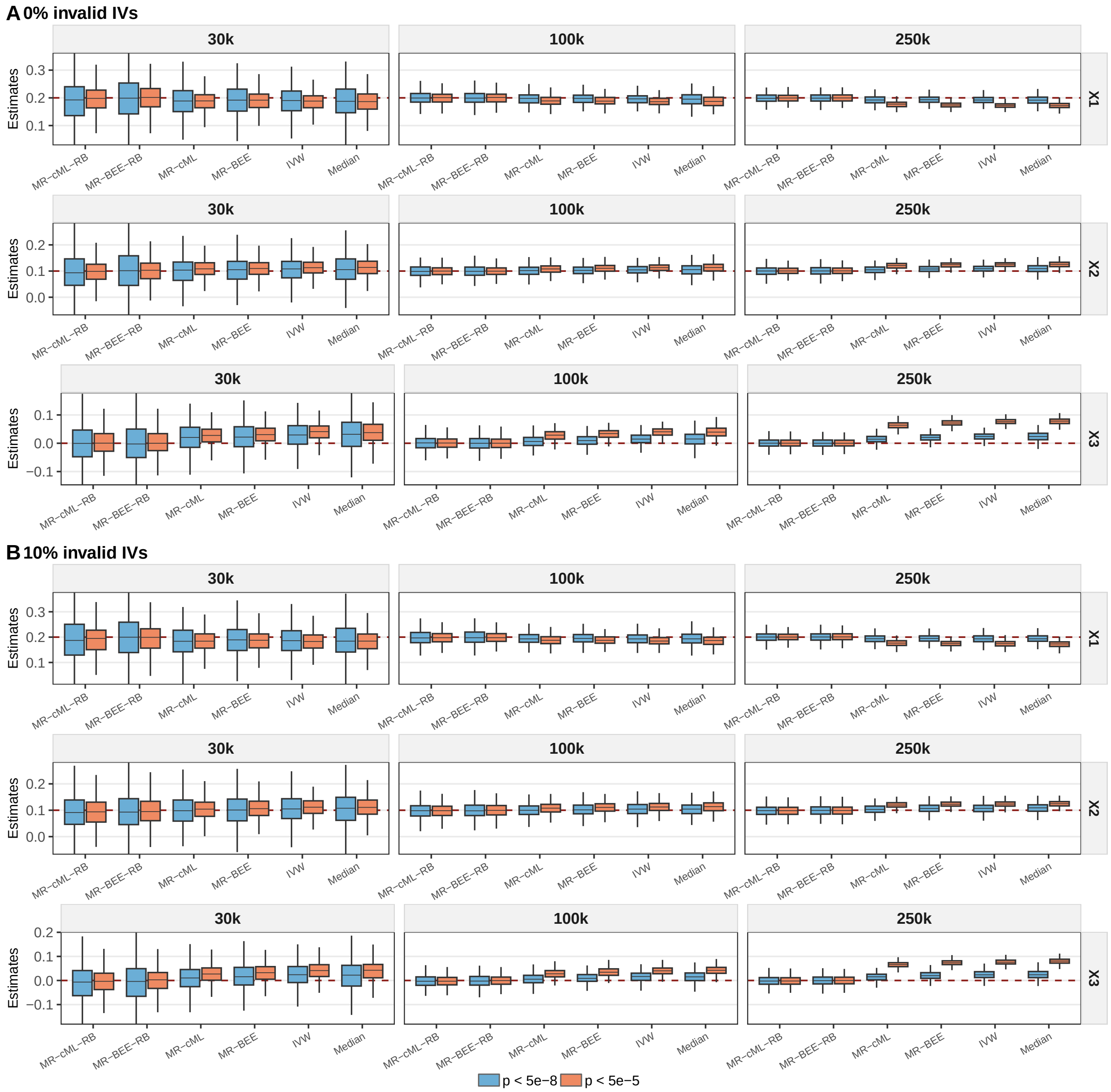

Figure S7: Boxplots of MVMR estimates in the winner's-curse + sample overlap + polygenicity setting. Rows corresponded to $X_1$, $X_2$, and $X_3$, and columns corresponded to sample sizes $N = 50{,}000$, 100,000, and 250,000. Within each panel, results were shown for IV selection thresholds $5 \times 10^{-8}$ and $5 \times 10^{-5}$.

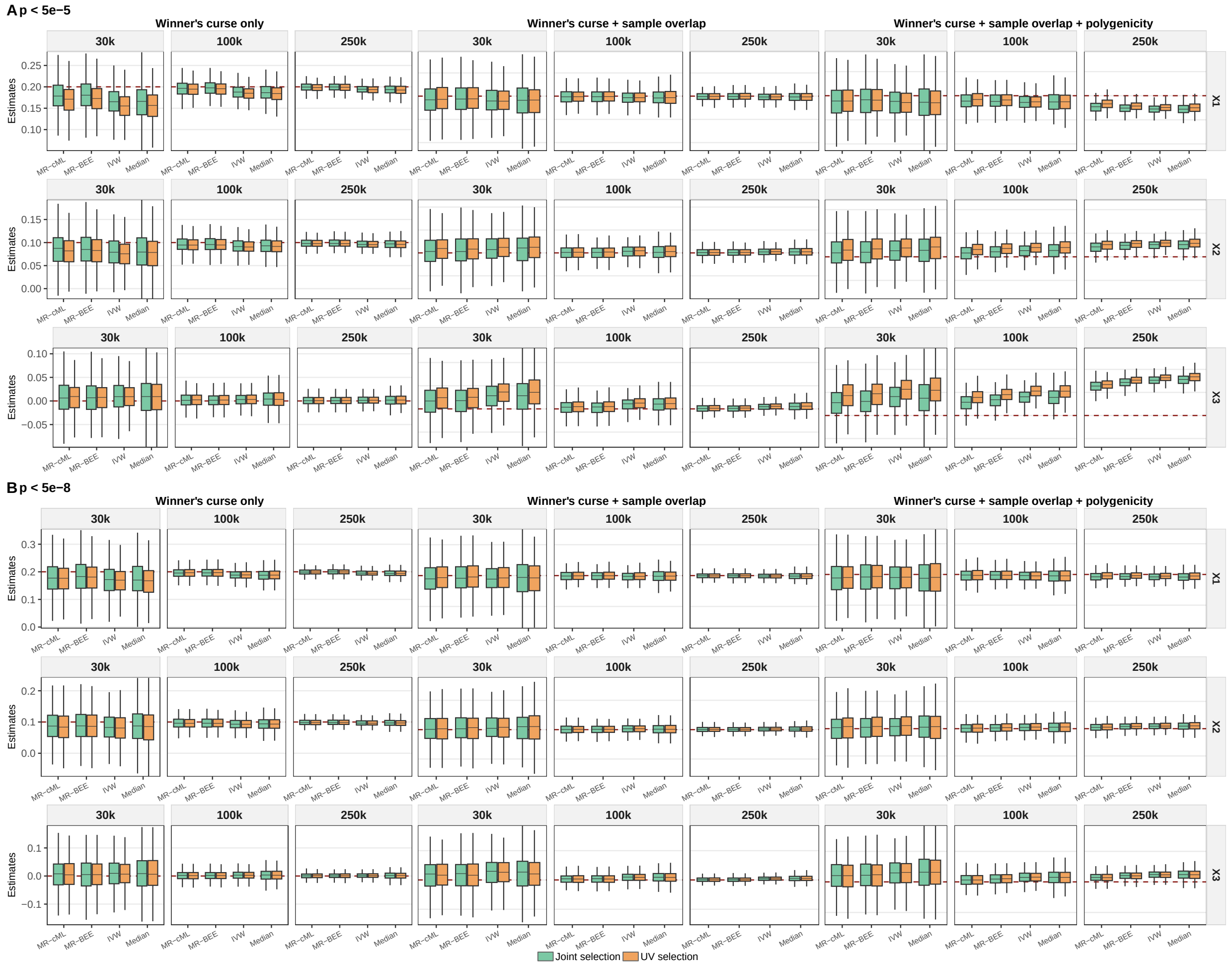

Figure S8: Comparison of multivariable MR estimates obtained using two IV selection strategies. Panels **A** and **B** show results under the thresholds $p < 5 \times 10^{-5}$ and $p < 5 \times 10^{-8}$, respectively.

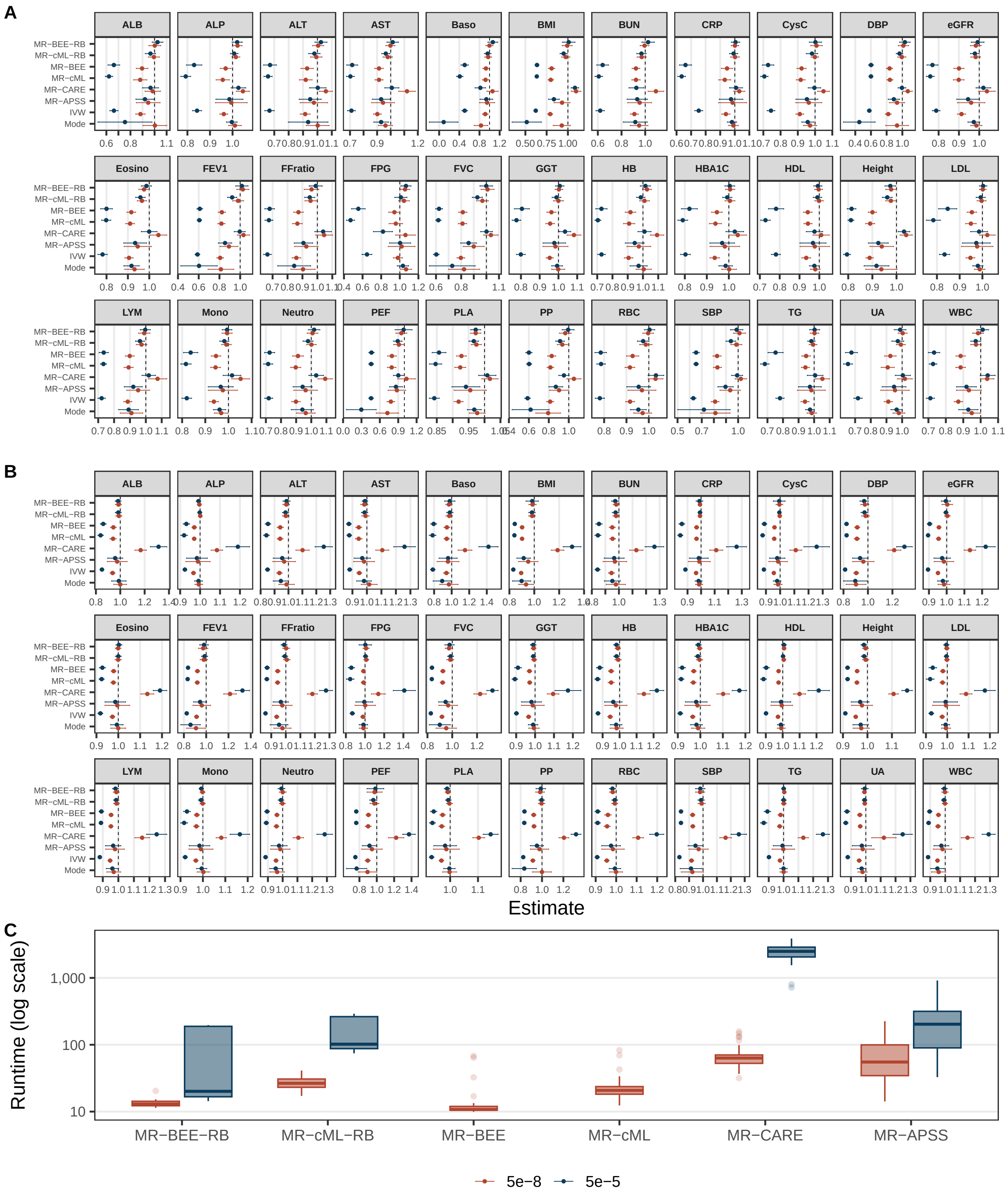

Figure S9: Causal effect estimates and computational runtime from the same-trait analysis. Causal effect estimates and 95% confidence intervals in **(A)** 0% overlap setting, and **(B)** 50% sample overlap setting. In both panels, results are shown under IV selection thresholds of $5 \times 10^{-5}$ (blue) and $5 \times 10^{-8}$ (red). **(C)** Boxplots of total runtime in seconds across the 33 traits in the 0% overlap setting. Runtime for each method includes LD pruning of IVs and estimation.

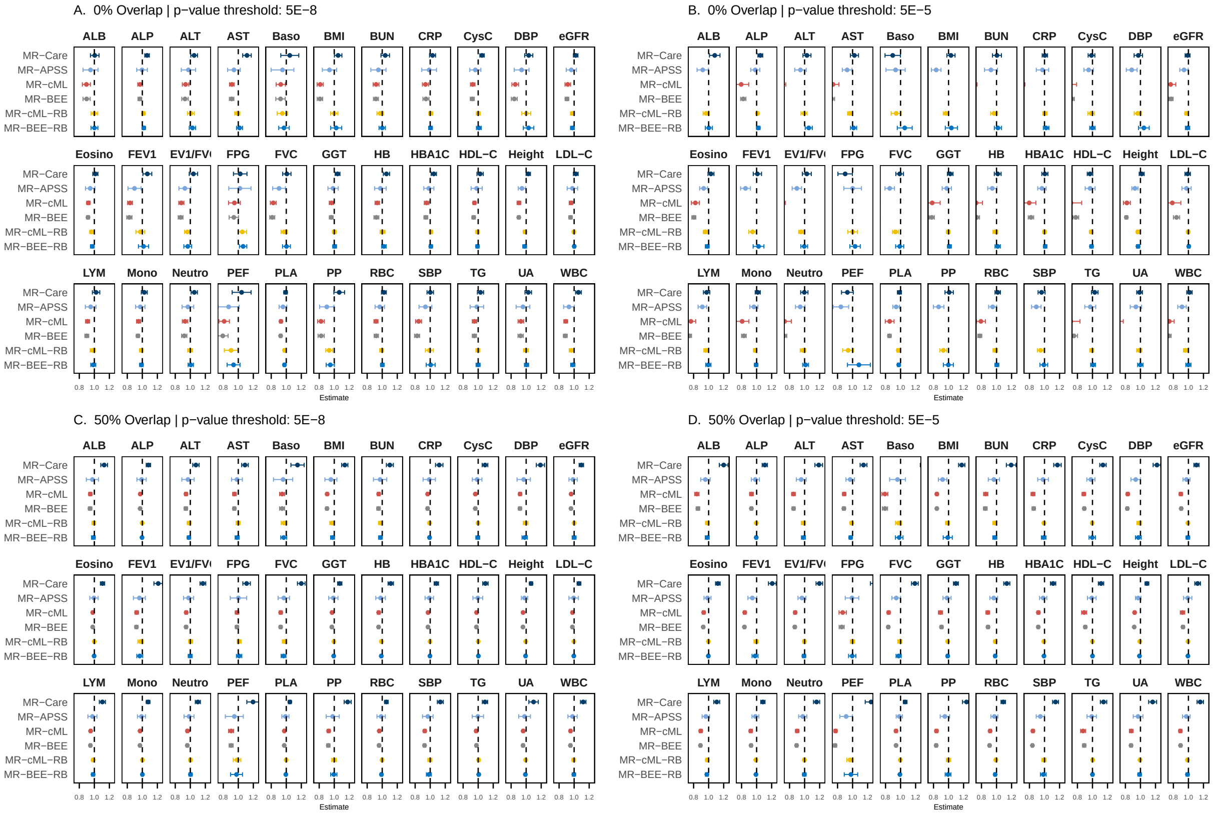

Figure S10: Robustness and independent replication of UVMR positive control benchmark results. Boxplots of causal effect estimates and nominal 95% confidence intervals from the same-trait analysis in both the 0% overlap and 50% setting across 33 UK Biobank quantitative traits. Results are shown under IV selection thresholds of $5 \times 10^{-5}$ and $5 \times 10^{-8}$. The dashed horizontal line indicates the target value of 1.

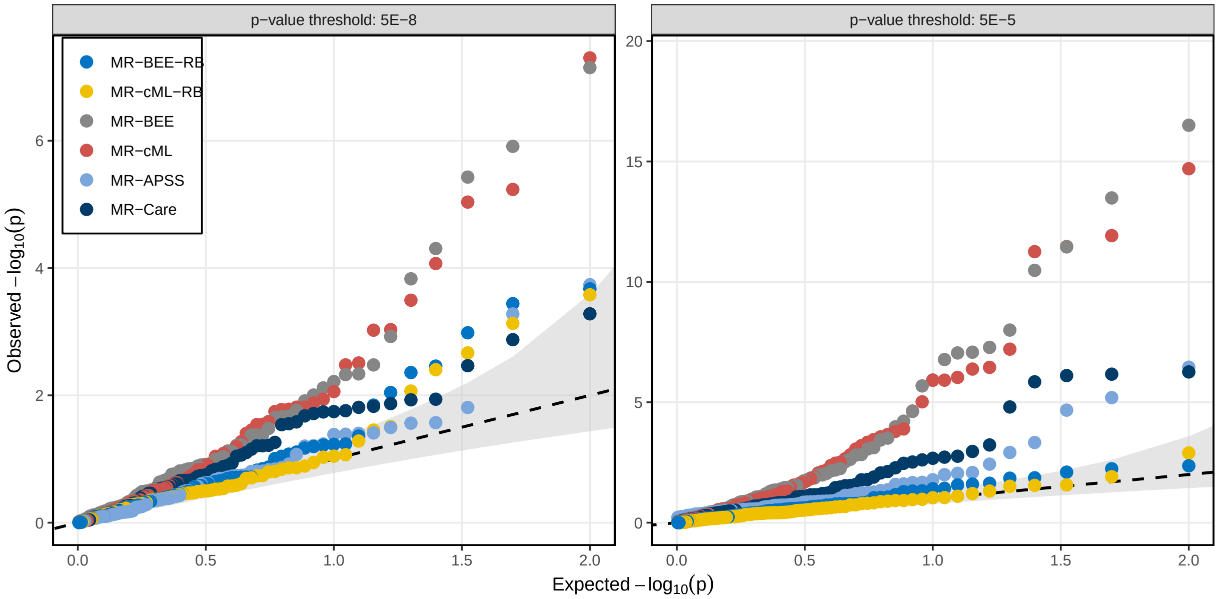

Figure S11: Robustness and independent replication of UVMR negative control benchmark results. QQ plots of $-\log_{10}(p)$ values. The gray-shaded part is 95% confidence interval. Results are shown under IV selection thresholds of $5 \times 10^{-5}$ (left) and $5 \times 10^{-8}$ (right).

### Supplemental Tables

Table S1: Definitions of phenotypes.

| Phenotype | Abbreviation | Data Field/Source |
|---|---|---|
| Cholesterol lowering medication | — | Data Fields 6153, 6177 |
| Blood pressure medication | — | Data Fields 6153, 6177 |
| Sex | Sex | Data Field 31 |
| Age | Age | Visit date − Birth date |
| Height | HEG | Data Field 50 |
| Body mass index | BMI | Data Field 21001 |
| Systolic blood pressure | SBP | Data Fields 93 and 4079 |
| Diastolic blood pressure | DBP | Data Fields 93 and 4079 |
| Pulse pressure | PP | SBP − DBP |
| Albumin | ALB | Data Field 30600 |
| Alkaline phosphatase | ALP | Data Field 30610 |
| Alanine aminotransferase | ALT | Data Field 30620 |
| Aspartate aminotransferase | AST | Data Field 30650 |
| Blood urea nitrogen | BUN | Data Field 30670 |
| Calcium | CA | Data Field 30680 |
| C-reactive protein | CRP | Data Field 30710 |
| Cystatin C | CysC | Data Field 30720 |
| Fasting plasma glucose | FPG | Data Field 30740 |
| Gamma-glutamyltransferase | GGT | Data Field 30730 |
| Glycated haemoglobin | HbA1c | Data Field 30750 |
| HDL cholesterol | HDL-C | Data Field 30760 |
| LDL cholesterol | LDL-C | Data Field 30780 |
| Triglycerides | TG | Data Field 30870 |
| Urate | UA | Data Field 30880 |
| White blood cell count | WBC | Data Field 30000 |
| Red blood cell count | RBC | Data Field 30010 |
| Platelet count | PLA | Data Field 30080 |
| Haemoglobin concentration | HB | Data Field 30030 |
| Monocyte count | Mono | Data Field 30120 |
| Lymphocyte count | LYM | Data Field 30110 |
| Neutrophil count | Neutro | Data Field 30100 |
| Forced vital capacity | FVC | Data Field 3062 |
| Forced expiratory volume in 1 s | FEV1 | Data Field 3063 |
| Peak expiratory flow | PEF | Data Field 3064 |
| Hair colour before greying - Dark brown | – | GCST90691600 |
| Hair colour before greying - Black | – | GCST90691601 |
| Coriander liking | – | GCST90094744 |

Table S2: GWAS summary data used in MVMR analysis.

| Trait | Abbr. | Type | Cohort 1 | | Cohort 2 | | Cohort 3 | |
|---|---|---|---|---|---|---|---|---|
| | | | Sample Size | PMID | Sample Size | PMID | Sample Size | PMID |
| Coronary artery disease | CAD | Binary | 210842/1167328 | 36474045 | 52789/262879 | 39024449 | 63307/416171 | r12.finngen.fi |
| Type 2 diabetes | T2D | Binary | 242283/1569734 | 38374256 | — | — | — | — |
| Smoking initiation | SMK | Binary | 801837 (total) | 36477530 | — | — | — | — |
| Drinks per week | DRNK | Continuous | 664400 | 36477530 | — | — | — | — |
| Body mass index | BMI | Continuous | 359983 | Neale lab | 424231 | 39024449 | 362327 | r12.finngen.fi |
| HDL cholesterol | HDL-C | Continuous | 1320016 | 34887591 | — | — | — | — |
| LDL cholesterol | LDL-C | Continuous | 1320016 | 34887591 | — | — | — | — |
| Systolic blood pressure | SBP | Continuous | 1028980 | 38689001 | — | — | — | — |
| Triglycerides | TG | Continuous | 1320016 | 34887591 | — | — | — | — |
| Urate | UA | Continuous | 354368 | 33462484 | — | — | — | — |

### Supplemental Notes

### S1 Rao-Blackwellization Theory

We establish the properties of the analytical Rao-Blackwellization in four steps. The conditions used are: (a) $\hat{\boldsymbol{\delta}}_j \sim \mathcal{N}(\boldsymbol{\delta}_j, \boldsymbol{\Sigma}_{\delta_j\delta_j})$ asymptotically, (b) $\boldsymbol{Z}_{\delta_j} \sim \mathcal{N}(\mathbf{0}, \eta^2\boldsymbol{\Sigma}_{\delta_j\delta_j})$ is generated independently of $\hat{\boldsymbol{\delta}}_j$, and (c) $\boldsymbol{\Sigma}_{\beta_j\beta_j} \succ 0$.

#### S1.1 Step 1: Noncentral chi-square distribution of the selection statistic.

The selection statistic conditional on $\hat{\boldsymbol{\delta}}_j$ follows a noncentral chi-square distribution.

*Proof.* By condition (b), $\hat{\boldsymbol{\beta}}_j + \boldsymbol{Z}_{\beta_j} \mid \hat{\boldsymbol{\delta}}_j \sim \mathcal{N}(\hat{\boldsymbol{\beta}}_j, \eta^2\boldsymbol{\Sigma}_{\beta_j\beta_j})$. By condition (c), perform the Cholesky decomposition $\boldsymbol{\Sigma}_{\beta_j\beta_j} = \mathbf{L}_j\mathbf{L}_j^\top$ and define

$$\boldsymbol{Y}_j = \frac{1}{\eta}\mathbf{L}_j^{-1}(\hat{\boldsymbol{\beta}}_j + \boldsymbol{Z}_{\beta_j}) \mid \hat{\boldsymbol{\delta}}_j \sim \mathcal{N}\left(\frac{1}{\eta}\mathbf{L}_j^{-1}\hat{\boldsymbol{\beta}}_j, \mathbf{I}_p\right).$$

Therefore,

$$S_j^{\mathrm{RB}}/\eta^2 \mid \hat{\boldsymbol{\delta}}_j = \|\boldsymbol{Y}_j\|^2 \mid \hat{\boldsymbol{\delta}}_j \sim \chi_p^2(\kappa_j),$$

where $\kappa_j = \eta^{-2}\|\mathbf{L}_j^{-1}\hat{\boldsymbol{\beta}}_j\|^2 = \eta^{-2}\hat{\boldsymbol{\beta}}_j^\top\boldsymbol{\Sigma}_{\beta_j\beta_j}^{-1}\hat{\boldsymbol{\beta}}_j$. □

#### S1.2 Step 2: Conditional moments of the exposure component.

We derive the conditional moments of $\boldsymbol{Z}_{\beta_j}$ given selection.

**Lemma 1** (Conditional moments under quadratic selection)**.** *Let $\boldsymbol{W} \sim \mathcal{N}(\mathbf{0}, \eta^2\mathbf{I}_p)$ and $\boldsymbol{\mu} \in \mathbb{R}^p$ with $\boldsymbol{\mu} \neq \mathbf{0}$. Define the selection event $A = \{\|\boldsymbol{\mu} + \boldsymbol{W}\|^2 > t\}$ with $P = \Pr(A) = \Pr(\chi_p^2(\|\boldsymbol{\mu}\|^2/\eta^2) > t/\eta^2) > 0$. Let $g_1 = \frac{\partial}{\partial\kappa}\log P$ and $g_2 = \frac{\partial^2}{\partial\kappa^2}\log P$ where $\kappa = \|\boldsymbol{\mu}\|^2/\eta^2$. Then*

$$\mathrm{E}[\boldsymbol{W} \mid A] = 2g_1\boldsymbol{\mu},$$
$$\mathrm{Cov}[\boldsymbol{W} \mid A] = \eta^2(1 + 2g_1)\mathbf{I}_p + 4g_2\boldsymbol{\mu}\boldsymbol{\mu}^\top.$$

*Proof.* By the rotational symmetry of $\boldsymbol{W}$ and of $A$ about the axis $\hat{\boldsymbol{\mu}} = \boldsymbol{\mu}/\|\boldsymbol{\mu}\|$, the conditional mean must be proportional to $\boldsymbol{\mu}$, $\mathrm{E}[\boldsymbol{W} \mid A] = c\,\boldsymbol{\mu}$, and the conditional covariance must take the form $\mathrm{Cov}[\boldsymbol{W} \mid A] = a\,\mathbf{I}_p + b\,\hat{\boldsymbol{\mu}}\hat{\boldsymbol{\mu}}^\top$ for scalars $c, a, b$.

To identify these scalars we use Stein-type identities for the noncentral chi-square. Set $\boldsymbol{Y} = \boldsymbol{\mu} + \boldsymbol{W} \sim \mathcal{N}(\boldsymbol{\mu}, \eta^2\mathbf{I}_p)$, so that $\|\boldsymbol{Y}\|^2/\eta^2 \sim \chi_p^2(\kappa)$ and $A = \{\|\boldsymbol{Y}\|^2 > t\}$. The survival function $P(\kappa)$ satisfies the differentiation identities

$$\frac{\partial}{\partial\kappa}\log P = g_1, \qquad \frac{\partial^2}{\partial\kappa^2}\log P = g_2,$$

and the tail moments of the noncentral chi-square give

$$\mathrm{E}[\|\boldsymbol{Y}\|^2/\eta^2 \mid A] = \kappa + p + 2\kappa g_1.$$

Expanding $\|\boldsymbol{Y}\|^2 = \|\boldsymbol{\mu}\|^2 + 2\boldsymbol{\mu}^\top\boldsymbol{W} + \|\boldsymbol{W}\|^2$ and matching with $\mathrm{E}[\boldsymbol{W} \mid A] = c\boldsymbol{\mu}$ yields $c = g_1$, hence $\mathrm{E}[\boldsymbol{W} \mid A] = 2g_1\boldsymbol{\mu}$.

For the covariance, applying the multivariate Stein identity $\mathrm{E}[h(\|\boldsymbol{Y}\|^2)\boldsymbol{Y}] = \boldsymbol{\mu}\,\mathrm{E}[h(\|\boldsymbol{Y}\|^2)] + 2\eta^2\,\mathrm{E}[h'(\|\boldsymbol{Y}\|^2)\boldsymbol{Y}]$ with $h(s) = \mathbf{1}[s > t]/P$, and differentiating $\log P$ with respect to $\kappa$ once and twice, identifies the two scalars as $a = \eta^2(1 + 2g_1)$ and $b\,\hat{\boldsymbol{\mu}}\hat{\boldsymbol{\mu}}^\top = 4g_2\boldsymbol{\mu}\boldsymbol{\mu}^\top$. Substituting gives the stated covariance. □

Applying Lemma 1 with $\boldsymbol{W} = \boldsymbol{Z}_{\beta_j}$ and $\boldsymbol{\mu} = \hat{\boldsymbol{\beta}}_j$ yields the exposure block of the conditional mean and covariance reported in the main text.

#### S1.3 Step 3: Conditional moments of the outcome component via Gaussian conditioning.

Since the outcome noise enters neither the selection statistic nor the selection event, its conditional moments follow from standard Gaussian conditioning.

**Lemma 2** (Gaussian conditioning under an exposure-only selection event)**.** *Let $(\boldsymbol{X}, \boldsymbol{Y})$ be jointly Gaussian with $\boldsymbol{X} \sim \mathcal{N}(\boldsymbol{\mu}_X, \boldsymbol{\Sigma}_{XX})$, $\boldsymbol{Y} \sim \mathcal{N}(\boldsymbol{\mu}_Y, \boldsymbol{\Sigma}_{YY})$, and cross-covariance $\boldsymbol{\Sigma}_{XY}$. For any event $A$ that depends only on $\boldsymbol{X}$,*

$$\begin{aligned}
\mathrm{E}[\boldsymbol{Y} \mid A] &= \boldsymbol{\mu}_Y + \boldsymbol{\Sigma}_{YX}\boldsymbol{\Sigma}_{XX}^{-1}(\mathrm{E}[\boldsymbol{X} \mid A] - \boldsymbol{\mu}_X),\\
\mathrm{Cov}[\boldsymbol{Y} \mid A] &= \boldsymbol{\Sigma}_{YX}\boldsymbol{\Sigma}_{XX}^{-1}\mathrm{Cov}[\boldsymbol{X} \mid A]\boldsymbol{\Sigma}_{XX}^{-1}\boldsymbol{\Sigma}_{XY} + \Big(\boldsymbol{\Sigma}_{YY} - \boldsymbol{\Sigma}_{YX}\boldsymbol{\Sigma}_{XX}^{-1}\boldsymbol{\Sigma}_{XY}\Big).
\end{aligned}$$

*Proof.* Write the Gaussian decomposition $\boldsymbol{Y} = \boldsymbol{\mu}_Y + \boldsymbol{\Sigma}_{YX}\boldsymbol{\Sigma}_{XX}^{-1}(\boldsymbol{X} - \boldsymbol{\mu}_X) + \boldsymbol{\varepsilon}$, where $\boldsymbol{\varepsilon} \sim \mathcal{N}(\mathbf{0}, \boldsymbol{\Sigma}_{YY} - \boldsymbol{\Sigma}_{YX}\boldsymbol{\Sigma}_{XX}^{-1}\boldsymbol{\Sigma}_{XY})$ is independent of $\boldsymbol{X}$. Since $A$ depends only on $\boldsymbol{X}$, $\boldsymbol{\varepsilon}$ remains independent of $A$ with unchanged moments under conditioning on $A$. Taking the conditional mean and covariance of both sides gives the result. □

Applying Lemma 2 with $\boldsymbol{X} = \boldsymbol{Z}_{\beta_j}$, $\boldsymbol{Y} = Z_{\alpha_j}$, $A = \{S_j^{\mathrm{RB}} > \lambda\}$, and the exposure moments from Step 2 yields the outcome block of the conditional moments reported in the main text.

#### S1.4 Step 4: Properties of the RB estimator.

**Property (i): Selection independence of the initial estimator.** The initial estimator $\hat{\boldsymbol{\delta}}_j^{\mathrm{init}} = \hat{\boldsymbol{\delta}}_j - \eta^{-2}\boldsymbol{Z}_{\delta_j}$ is uncorrelated with the randomized exposure statistic,

$$\mathrm{cov}\Big(\hat{\boldsymbol{\delta}}_j^{\mathrm{init}}, \hat{\boldsymbol{\beta}}_j + \boldsymbol{Z}_{\beta_j}\Big) = \mathrm{cov}(\hat{\boldsymbol{\delta}}_j, \hat{\boldsymbol{\beta}}_j) - \eta^{-2}\mathrm{cov}(\boldsymbol{Z}_{\delta_j}, \boldsymbol{Z}_{\beta_j}) = \mathbf{0},$$

where the cancellation uses $\mathrm{cov}(\boldsymbol{Z}_{\delta_j}, \boldsymbol{Z}_{\beta_j}) = \eta^2\boldsymbol{\Sigma}_{\delta_j\beta_j}$ and $\mathrm{cov}(\hat{\boldsymbol{\delta}}_j, \hat{\boldsymbol{\beta}}_j) = \boldsymbol{\Sigma}_{\delta_j\beta_j}$. Under the joint Gaussianity of $(\hat{\boldsymbol{\delta}}_j, \boldsymbol{Z}_{\delta_j})$, zero covariance implies independence. Since $S_j^{\mathrm{RB}}$ is a function of $\hat{\boldsymbol{\beta}}_j + \boldsymbol{Z}_{\beta_j}$ alone, we conclude $\hat{\boldsymbol{\delta}}_j^{\mathrm{init}} \perp \mathbb{I}(S_j^{\mathrm{RB}} > \lambda)$.

**Property (ii): Conditional unbiasedness.** By the tower property, the definition $\hat{\boldsymbol{\delta}}_j^{\mathrm{RB}} = \mathrm{E}[\hat{\boldsymbol{\delta}}_j^{\mathrm{init}} \mid \hat{\boldsymbol{\delta}}_j, S_j^{\mathrm{RB}} > \lambda]$, and Property (i),

$$\begin{aligned}\mathrm{E}[\hat{\boldsymbol{\delta}}_j^{\mathrm{RB}} \mid S_j^{\mathrm{RB}} > \lambda] &= \mathrm{E}\Big[\mathrm{E}[\hat{\boldsymbol{\delta}}_j^{\mathrm{init}} \mid \hat{\boldsymbol{\delta}}_j,\, S_j^{\mathrm{RB}} > \lambda] \,\Big|\, S_j^{\mathrm{RB}} > \lambda\Big] \\ &= \mathrm{E}[\hat{\boldsymbol{\delta}}_j^{\mathrm{init}} \mid S_j^{\mathrm{RB}} > \lambda] = \mathrm{E}[\hat{\boldsymbol{\delta}}_j^{\mathrm{init}}] = \boldsymbol{\delta}_j,\end{aligned}$$

where the third equality uses the independence in Property (i) and the last uses $\mathrm{E}[\hat{\boldsymbol{\delta}}_j^{\mathrm{init}}] = \mathrm{E}[\hat{\boldsymbol{\delta}}_j] = \boldsymbol{\delta}_j$.

**Property (iii): Conditional covariance and its plug-in estimator.** By the conditional variance decomposition, conditioning throughout on the selection event $S_j^{\mathrm{RB}} > \lambda$,

$$\mathrm{Cov}(\hat{\boldsymbol{\delta}}_j^{\mathrm{init}} \mid S_j^{\mathrm{RB}} > \lambda) = \mathrm{Cov}(\hat{\boldsymbol{\delta}}_j^{\mathrm{RB}} \mid S_j^{\mathrm{RB}} > \lambda) + \mathrm{E}\Big[\mathrm{Cov}(\hat{\boldsymbol{\delta}}_j^{\mathrm{init}} \mid \hat{\boldsymbol{\delta}}_j,\, S_j^{\mathrm{RB}} > \lambda) \,\Big|\, S_j^{\mathrm{RB}} > \lambda\Big].$$

By Property (i) the left-hand side equals the unconditional covariance $\mathrm{Cov}(\hat{\boldsymbol{\delta}}_j^{\mathrm{init}}) = (1 + \eta^{-2})\boldsymbol{\Sigma}_{\boldsymbol{\delta}_j\boldsymbol{\delta}_j}$. Since $\boldsymbol{Z}_{\boldsymbol{\delta}_j} \perp \hat{\boldsymbol{\delta}}_j$ by condition (b), the inner conditional covariance reduces to $\mathrm{Cov}(\hat{\boldsymbol{\delta}}_j^{\mathrm{init}} \mid \hat{\boldsymbol{\delta}}_j,\, S_j^{\mathrm{RB}} > \lambda) = \eta^{-4}\mathrm{Cov}(\boldsymbol{Z}_{\boldsymbol{\delta}_j} \mid \hat{\boldsymbol{\delta}}_j,\, S_j^{\mathrm{RB}} > \lambda)$. Rearranging gives the exact conditional covariance

$$\mathrm{Cov}(\hat{\boldsymbol{\delta}}_j^{\mathrm{RB}} \mid S_j^{\mathrm{RB}} > \lambda) = (1 + \eta^{-2})\boldsymbol{\Sigma}_{\boldsymbol{\delta}_j\boldsymbol{\delta}_j} - \eta^{-4}\,\mathrm{E}\Big[\mathrm{Cov}(\boldsymbol{Z}_{\boldsymbol{\delta}_j} \mid \hat{\boldsymbol{\delta}}_j,\, S_j^{\mathrm{RB}} > \lambda) \,\Big|\, S_j^{\mathrm{RB}} > \lambda\Big].$$

The outer expectation is taken over the post-selection distribution of $\hat{\boldsymbol{\delta}}_j$ and depends on the unknown $\boldsymbol{\delta}_j$. Following the plug-in strategy of Ma et al. (2023), we evaluate the inner conditional covariance at the observed $\hat{\boldsymbol{\delta}}_j$ and omit the outer expectation, defining

$$\hat{\boldsymbol{\Sigma}}_{\boldsymbol{\delta}_j\boldsymbol{\delta}_j}^{\mathrm{RB}} = (1 + \eta^{-2})\boldsymbol{\Sigma}_{\boldsymbol{\delta}_j\boldsymbol{\delta}_j} - \eta^{-4}\,\mathrm{Cov}(\boldsymbol{Z}_{\boldsymbol{\delta}_j} \mid \hat{\boldsymbol{\delta}}_j,\, S_j^{\mathrm{RB}} > \lambda).$$

By the law of iterated expectation, $\mathrm{E}[\hat{\boldsymbol{\Sigma}}_{\boldsymbol{\delta}_j\boldsymbol{\delta}_j}^{\mathrm{RB}} \mid S_j^{\mathrm{RB}} > \lambda] = \mathrm{Cov}(\hat{\boldsymbol{\delta}}_j^{\mathrm{RB}} \mid S_j^{\mathrm{RB}} > \lambda)$, so $\hat{\boldsymbol{\Sigma}}_{\boldsymbol{\delta}_j\boldsymbol{\delta}_j}^{\mathrm{RB}}$ is conditionally unbiased for the target covariance.

### S2 Additional simulation results

#### S2.1 UVMR with 10% invalid IVs

We additionally considered a UVMR setting with 10% invalid IVs. As in the main-text UV simulations, we generated sparse effects among $M = 50{,}000$ variants, with 1% of variants having $\beta_j \neq 0$. We used $q_{\mathrm{inv}} = 0.1$ to denote the proportion of invalid variants among these sparse signals. The valid IVs were then generated from a sparse subset with proportion $0.01(1 - q_{\mathrm{inv}})$, while the invalid IVs were divided equally into two additional sparse subsets, each with proportion $0.01q_{\mathrm{inv}}/2$. For the first half of invalid IVs, variants had direct effects $\gamma_j \sim \mathcal{N}(0, 0.01^2)$. For the second half, the IV strength ($\beta_j$) and direct effects ($\gamma_j$) were jointly generated from a bivariate normal distribution to represent correlated pleiotropy. The nonzero exposure effects were generated as in the main text.

To compare with the main-text UV results, Figure S1 shows the boxplots under the main text setting $q_{\text{inv}} = 0$ for both $\theta = 0$ and $\theta = 0.1$, while Figure S2 shows the corresponding results for the setting $q_{\text{inv}} = 0.1$. In Figure S1, when there were no invalid IVs and no background noise, MR-BEE-RB, MR-cML-RB, MR-CARE, and MR-APSS were unbiased, while methods that did not correct for winner's curse showed the expected mild attenuation, and this bias became smaller as $N$ increased. In Figure S2, after adding 10% invalid IVs but still keeping no background noise, MR-APSS was clearly biased, likely because its foreground model did not accommodate correlated pleiotropy. IVW was also biased even at large $N$, which was consistent with violation of the InSIDE assumption. By contrast, MR-BEE-RB, MR-cML-RB, and MR-CARE remained essentially unbiased, and the non-RB MR-BEE and MR-cML also became close to unbiased once $N$ was large enough that winner's-curse bias was small. Once background noise was introduced, all methods except MR-BEE-RB and MR-cML-RB were biased. Figure S3 showed the same overall pattern for coverage as in the main text. MR-BEE-RB, MR-cML-RB, MR-APSS and weighted mode stayed closest to nominal across all settings, while other methods showed increasing undercoverage with sample overlap and especially the sample overlap + polygenicity setting.

#### S2.2 Additional MVMR scenarios

We next considered several additional MVMR scenarios. As in the main-text MV simulations, we generated sparse multivariable exposure effects among $M = 50{,}000$ variants, with 1% of variants having nonzero effects under a mixed architecture that allowed a variant to affect one, two, or all three exposures. We used $q_{\text{inv}}$ to denote the proportion of invalid variants among these sparse signals. When $q_{\text{inv}} = 0.1$, 10% of the sparse signals were treated as invalid IVs and additionally had direct effects $\gamma_j \sim \mathcal{N}(0, 0.01^2)$, while the remaining sparse signals were valid IVs. The nonzero exposure effects were generated as in the main text.

##### S2.2.1 Winner's curse only

We considered a winner's-curse-only scenario in the MVMR setting. This scenario includes both 0% and 10% invalid IV settings and follows the same MV framework described above. In this case, $\mathbf{R}_{\varepsilon\varepsilon} = \mathbf{I}_4$ and $\boldsymbol{\Omega}_j = \mathbf{0}$.

Figure S4 showed that, with 0% invalid IVs, methods that did not correct for winner's curse showed the expected slight attenuation towards the null, especially at $N = 30{,}000$ and under $5 \times 10^{-5}$, and this became smaller as $N$ increased. The same overall pattern remained after adding 10% invalid IVs. MR-BEE-RB and MR-cML-RB stayed unbiased across $X_1$, $X_2$, and $X_3$, while MR-BEE and MR-cML also became nearly unbiased once $N$ was moderate to large. IVW and weighted median still showed the largest attenuation. Coverage stayed close to 95% overall (Figure S6).

##### S2.2.2 Winner's curse plus sample overlap

We considered a scenario with sample overlap but no polygenicity in the MVMR setting. This scenario includes both 0% and 10% invalid IV settings and follows the same MV framework described above. Here

$$\mathbf{R}_{\varepsilon\varepsilon} = \begin{pmatrix} 1 & 0.2 & 0.2 & 0.2 \\ 0.2 & 1 & 0.2 & 0.2 \\ 0.2 & 0.2 & 1 & 0.2 \\ 0.2 & 0.2 & 0.2 & 1 \end{pmatrix}$$

and $\mathbf{\Omega}_j = \mathbf{0}$.

Figure S5 showed that sample overlap induced a more complicated bias pattern in MVMR than winner's curse alone. For $X_1$, which had the largest true effect, the non-RB methods tended to show downward bias, whereas for $X_2$ and $X_3$ the bias was generally upward. This bias was mild for MR-BEE and MR-cML, but more pronounced for IVW and Median, especially at $N = 30{,}000$ and under $5 \times 10^{-5}$, and it became smaller as $N$ increased. By contrast, MR-BEE-RB and MR-cML-RB remained close to unbiased across $X_1$, $X_2$, and $X_3$ in both the 0% and 10% invalid-IV settings. Coverage was still fairly close to 95% overall (Figure S6).

##### S2.2.3 Winner's curse plus sample overlap and polygenicity

In the scenario with both sample overlap and polygenicity, the covariance structure used

$$\mathbf{R}_{\varepsilon\varepsilon} = \begin{pmatrix} 1 & 0.2 & 0.2 & 0.2 \\ 0.2 & 1 & 0.2 & 0.2 \\ 0.2 & 0.2 & 1 & 0.2 \\ 0.2 & 0.2 & 0.2 & 1 \end{pmatrix}$$

and

$$\mathbf{\Omega}_j = \frac{1}{50{,}000} \begin{pmatrix} 0.3 & 0.06 & 0.06 & 0.06 \\ 0.06 & 0.3 & 0.06 & 0.06 \\ 0.06 & 0.06 & 0.3 & 0.06 \\ 0.06 & 0.06 & 0.06 & 0.3 \end{pmatrix},$$

with rows and columns corresponding to $(X_1, X_2, X_3, Y)$.

Figures S7 and S6 showed that the main-text MV pattern persisted after adding 10% invalid IVs. MR-BEE-RB and MR-cML-RB stayed almost unbiased across $X_1$, $X_2$, and $X_3$, under both thresholds and all three sample sizes. By contrast, MR-BEE, MR-cML, IVW, and weighted median were downward biased for $X_1$ and upward biased for $X_2$ and $X_3$, and that bias was more pronounced under the more liberal threshold $5 \times 10^{-5}$ and at larger $N$. The two RB methods also stayed closest to 95% coverage, while the other methods undercovered, with the worst deterioration again showing up at $5 \times 10^{-5}$ and $N = 250{,}000$.

##### S2.2.4 Comparison between IV selection strategies

We also compared two common IV selection strategies used in the MVMR literature. The first was a marginal UV-then-union strategy, in which IVs were selected separately for each exposure based on the univariable association statistics and the resulting IV sets were combined by union. The second was a multivariable joint-selection strategy based on the joint chi-square test statistic that evaluates whether a variant is associated with at least one exposure. We applied both strategies to MR-cML, MR-BEE, IVW and weighted median to assess how the choice of IV selection affected finite-sample bias.

As shown in Figure S8, the two strategies gave very similar results under the stringent threshold $5 \times 10^{-8}$, where both procedures selected relatively strong instruments. Under the more liberal threshold $5 \times 10^{-5}$, however, the UV-then-union strategy was generally more biased than the joint-selection strategy. This was likely because the UV-then-union strategy tended to select more weak instruments that were more susceptible to winner's curse and polygenicity bias, whereas the joint-selection strategy tended to select fewer but stronger instruments.

### S3 Real data application details

#### S3.1 GWAS summary statistics

For the UV analysis, we used UKB participants of genetically inferred (Privé et al. 2022) and self-reported European ancestry (Data Field: 21000), after removing related individuals (kinship $> 0.0884$) and withdrawn participants. For the 0% sample overlap setting, we generated exposure and outcome GWAS summary statistics by randomly splitting the full sample into two independent groups stratified by age bins and sex. For the 50% sample overlap setting, we instead split the full sample into three parts, with one part serving as the shared component. Medication status (Data Field: 6153, 6177) was adjusted for LDL cholesterol, diastolic blood pressure (DBP), and systolic blood pressure (SBP), following standard procedures used in the literature (Graham et al. 2021, Keaton et al. 2024). In the positive control analysis, we used both the 0% and 50% overlap GWAS configurations. In the negative control analysis, we obtained the full-sample GWAS summary statistics for the negative outcome traits by combining the two non-overlapping GWAS results from the 0% overlap setting via inverse variance weighting meta-analysis. All GWAS summary statistics used in the UV analyses were harmonized to the same reference panel of 7.35 million SNPs imputed using SBayesRC (Zheng et al. 2024). We computed LD scores from the same reference panel, and the resulting LDSC-derived quantities were used in downstream methods that accounted for sample overlap and background structure. Table S1 summarizes the GWASs used in UVMR analysis.

For the MV analysis, we obtained GWAS summary statistics for CAD from (Aragam et al. 2022). The ten exposures — body mass index (BMI), smoking initiation (SMK), drinks per week (DRNK), DBP, SBP, HDL cholesterol, LDL cholesterol, TG, T2D, and urate (UA) — are established cardiometabolic risk factors for CAD, with GWAS summary statistics sourced from (Graham et al. 2021, Saunders et al. 2022, Keaton et al. 2024, Verma et al. 2024, Smith et al. 2024). All summary statistics were harmonized to a reference panel of 7.35 million SNPs imputed using SBayesRC (Zheng et al. 2024), and LDSC was used to estimate the background noise structure. Table S2 summarizes the GWASs used in MVMR analysis.

##### S3.2 IV selection

For the UV analysis, MR-cML-RB and MR-BEE-RB used rerandomized exposure-GWAS $p$-values with $\eta = 0.5$ for IV selection at thresholds $5 \times 10^{-8}$ and $5 \times 10^{-5}$, where the selection statistic accounted for sample overlap, polygenic background and GWAS inflation through LDSC-derived quantities. Then we performed clumping with PLINK using a 10,000 kb window and $r^2 < 0.001$, with 1,000 randomly selected UKB individuals as the LD reference panel. MR-CARE used its main function `mr_care()` to select IVs and perform clumping with the same LD reference panel and clumping parameters. The remaining methods used variants selected based on the raw exposure GWAS $p$-values at the corresponding threshold and performed LD clumping in PLINK with the same configuration.

For the MV analysis, MR-cML-RB and MR-BEE-RB used rerandomized multivariable joint $p$-values with $\eta = 0.5$ for IV selection, where the per-SNP covariance structure accounted for background noises. For comparison, the non-adjusted analysis used the original multivariable joint test without rerandomization and the background-signal correction. In each case, we performed PLINK clumping at joint $p < 5 \times 10^{-8}$ with window = 1,000 kb and $r^2 < 0.001$, which yielded 1,217 independent IVs for the adjusted analysis and 3,198 independent IVs for the non-adjusted analysis.

##### S3.3 Supplemental Results

To assess the reproducibility of the proposed framework under its randomized IV selection, we independently replicated the UVMR benchmark analyses twice using entirely different computational resources, with each implementation coded independently by different authors of this paper. Despite the inherent randomness of the IV selection step, the results were highly consistent across both implementations, as shown in Figures S10-S11.

#### S4 Implementation details and tuning parameters.

The Rao–Blackwellization step involves several numerical choices that are fixed a priori in our implementation. First, because the covariance components $\mathbf{R}_{\varepsilon\varepsilon}$ and $\boldsymbol{\Omega}$ are estimated from LDSC, all summary statistics are put on the same standardized scale before correction. In particular, the GWAS effect estimates are represented on the scale $\hat{\delta}_{js} = Z_{js}/\sqrt{n_s}$, with corresponding standard errors $1/\sqrt{n_s}$, or equivalently on the scale obtained after standardizing both phenotypes and genotypes. This ensures that

$$\boldsymbol{\Sigma}_{\boldsymbol{\delta}_j\boldsymbol{\delta}_j} = \mathbf{D}_j \mathbf{R}_{\varepsilon\varepsilon} \mathbf{D}_j + l_j \boldsymbol{\Omega}$$

has the same units as $\hat{\boldsymbol{\delta}}_j = (\hat{\boldsymbol{\beta}}_j^\top, \hat{\alpha}_j)^\top$.

The randomization scale is fixed at $\eta = 0.5$, following the rerandomized-selection literature and our empirical evaluation. This choice provides a stable compromise between staying close to the original selection rule and avoiding excessive dependence on the auxiliary randomization. We also exclude variants whose conditional selection probability

$$P_j = \Pr\left\{\chi_p^2(\kappa_j) > \lambda/\eta^2\right\}, \qquad \kappa_j = \eta^{-2} \hat{\boldsymbol{\beta}}_j^\top \boldsymbol{\Sigma}_{\beta_j\beta_j}^{-1} \hat{\boldsymbol{\beta}}_j,$$

is below $P_{\text{thres}} = 0.1$. Such variants are typically selected only under an unusually favorable realization of the auxiliary noise, making their conditional moments unstable. In addition, if the exposure block $\mathbf{\Sigma}_{\boldsymbol{\beta}_j\boldsymbol{\beta}_j}$ is not positive definite, the noncentral-$\chi^2$ calculation is not well defined and the variant is removed from the corrected IV set. Because the RB covariance is a plug-in estimate, small negative diagonal entries may occasionally arise from finite-sample or numerical error; the reported RB standard errors are therefore computed from the nonnegative part of the diagonal, and variants with zero corrected exposure variance are removed from downstream procedures requiring matrix inversion.

Finally, for traits or exposure dimensions with low heritability, fully removing estimation-error bias can lead to unstable estimating equations, a phenomenon also observed in measurement-error-corrected MR methods (Lorincz-Comi et al. 2024). This issue is especially relevant in MVMR, where a jointly selected IV may be strong for one exposure but weak for another. We therefore apply a partial covariance stabilization to the RB covariance matrix. For exposure $s$ of variant $j$, define the empirical reliability

$$\rho_{js} = \frac{(\hat{\beta}_{js}^{\text{RB}})^2 - \left(\hat{\mathbf{\Sigma}}_{\boldsymbol{\delta}_j\boldsymbol{\delta}_j}^{\text{RB}}\right)_{ss}}{(\hat{\beta}_{js}^{\text{RB}})^2}.$$

Let $\mathbf{L}_j$ be a diagonal matrix with $(\mathbf{L}_j)_{p+1,p+1} = 1$. For the exposure coordinates, we set

$$(\mathbf{L}_j)_{ss} = \begin{cases} 1, & \rho_{js} > r, \\ \left\{ \dfrac{(1-r)(\hat{\beta}_{js}^{\text{RB}})^2}{\left(\hat{\mathbf{\Sigma}}_{\boldsymbol{\delta}_j\boldsymbol{\delta}_j}^{\text{RB}}\right)_{ss}} \right\}^{1/2}, & \rho_{js} \le r, \end{cases}$$

and replace

$$\hat{\mathbf{\Sigma}}_{\boldsymbol{\delta}_j\boldsymbol{\delta}_j}^{\text{RB}} \leftarrow \mathbf{L}_j \hat{\mathbf{\Sigma}}_{\boldsymbol{\delta}_j\boldsymbol{\delta}_j}^{\text{RB}} \mathbf{L}_j.$$

This adjustment leaves reliable exposure dimensions unchanged, while shrinking the covariance correction for exposure-specific weak instruments. Larger values of $r$ impose stronger stabilization by shrinking more aggressively; as $r \to 1$, the corresponding covariance correction is nearly removed. We use $r = 0.3$ as the default, which provides stable estimates in low-heritability settings while retaining most of the estimation-error correction. This adjustment is first used in our work (Yang et al. 2024).

In routine applications, we recommend using the default values $\eta = 0.5$, $P_{\text{thres}} = 0.1$, and $r = 0.3$, and keeping them fixed across analyses.
